# The role of the components of PM_2.5_ in the incidence of Alzheimer’s disease and related disorders

**DOI:** 10.1101/2024.12.10.24318725

**Authors:** Haisu Zhang, Yifan Wang, Haomin Li, Qiao Zhu, Tszshan Ma, Yang Liu, Kyle Steenland

## Abstract

**Background:** The associations of PM_2.5_ mass and various adverse health outcomes have been widely investigated. However, fewer studies focused on the potential health impacts of PM_2.5_ components, especially for dementia and Alzheimer’s diseases (AD).

**Methods:** We constructed a nationwide population-based open cohort study among Medicare beneficiaries aged 65 or older during 2000-2018. This dataset was linked with the predicted levels of 15 PM_2.5_ components, including 5 major mass contributors (EC, OC, NH_4_^+^, NO_3_^-^, SO_4_^2-^) and 10 trace elements (Br, Ca, Cu, Fe, K, Ni, Pb, Si, V, Zn) across contiguous US territory. Data were aggregated by ZIP code, calendar year and individual level demographics. Two mixture analysis methods, weighted quantile sum regression (WQS) and quantile g-computation (qgcomp), were used with quasi-Poisson models to analyze the health effects of the total mixture of PM_2.5_ components on dementia and AD, as well as the relative contribution of individual components.

**Results:** Exposure to PM_2.5_ components over the previous 5 years was significantly associated with increased risks of both dementia and AD, with stronger associations observed for AD. SO_4_^2-^, OC, Cu were identified with large contributions to the combined positive association of the mixture from both WQS and qgcomp models.

**Conclusion:** We found positive associations between the 15 PM_2.5_ components and the incidence of dementia and AD. Our findings suggest that reducing PM_2.5_ emissions from traffic and fossil fuel combustion could help mitigate the growing burden of dementia and Alzheimer’s disease.

## Introduction

In 2019, the estimated number of dementia patients worldwide exceeded 55 million, with societal costs surpassing 1,300 billion US dollars (1). In the United States alone, dementia affected more than 6.7 million Americans in 2023, and projections indicate 14 million cases by 2060 (2). Alzheimer’s disease (AD) is the most common disease that leads to dementia and was the 6^th^ leading cause of death in the United States (3). Given the absence of a cure for AD and related dementias (ADRD), a promising strategy to alleviate the health burden associated with these conditions is the identification and control of potential modifiable risk factors.

Growing evidence supports a relationship between air pollution and dementia, particularly regarding particulate matter with diameters less than 2.5 micrometers (PM_2.5_). Epidemiological studies conducted globally have consistently reported a positive association between long-term exposure to PM_2.5_ mass and the risk of dementia (4-7).

The complex composition of PM_2.5_ can benefit from a detailed examination, as the pathogenic effects of PM_2.5_ are not precisely elucidated when treated as a whole. Health studies at the component level are needed to quantify the relative hazards of specific components, providing actionable recommendations to policymakers for targeted pollution control. Numerous investigations have revealed positive long-term and short-term associations between PM_2.5_ components and various health outcomes, including non-accidental mortality (8-10), cardiovascular diseases (11, 12), and respiratory diseases (13, 14). However, few studies have investigated the health effects of PM_2.5_ components on dementia using a mixture analysis approach.

Another important limitation of past studies on the health effects of PM_2.5_ components is the lack of high-resolution exposure data. While some previous studies have relied on data from ground-based monitoring stations (15, 16), such data often fails capture the potentially greater spatial heterogeneity of some components comparing to PM_2.5_ mass. Moreover, the relatively limited number of monitoring stations for specific PM_2.5_ components has limited the scope of these studies. To cope with these challenges, many studies started to use data from exposure prediction models as an alternative (8, 11). Such models can achieve higher spatial resolution and good prediction accuracies, allowing for better estimation of their associations with adverse health outcomes.

A significant challenge in exploring potential relationships between PM_2.5_ components and adverse health outcomes is the observed high correlation among different particle components, as documented in previous studies (15, 17, 18). Our previous study evaluated 5 major PM_2.5_ components (EC, OC, NH_4_^+^, NO_3_^-^, SO_4_^2-^) and showed positive associations with AD and ADRD through single pollutant models (19). Here, we extend that research to incorporate the high-resolution data of 15 PM_2.5_ components and utilize two mixture analysis methods, which can control for component correlations: weighted quantile sum regression and quantile g-computation, to derive more robust and comprehensive estimates on the relative toxicity of these components.

## Methods

### Study Population

Two nationwide, privacy-protected and publicly available databases from the Centers for Medicare and Medicaid Services (CMS), including the Medicare denominator file and the Medicare Chronic Conditions Warehouse (CCW), were analyzed in this study. The denominator file contains enrollment records for each Medicare beneficiary, including demographics, Medicaid insurance status (a proxy for SES), the date of death, and ZIP code of residence, which were updated annually. The CCW claims data include predefined indicators for chronic conditions, based on ICD codes from health care providers, among the fee-for-service (FFS) Medicare beneficiaries and provides the date of the first occurrence with a diagnosis code for a specific condition.

We constructed separate cohorts for all-cause dementia and AD with the records from 2000 to 2018. For each cohort, we further required a “clean” period of 5 years after enrollment, during which the eligible beneficiaries could not have diagnosis codes for dementia or AD. For example, a beneficiary entering Medicare in 2003 would be required to be dementia-free by 2008; and the follow-up would only begin in 2008. The removal of potentially prevalent cases during the first 5 years of follow up was done to estimate incidence rather than prevalence effect measures. Overall, the study subjects entered the cohort on January 1^st^ of the year following the “clean” period and were followed until the first diagnosis of the outcome of interest, death, or end of follow-up.. All subjects were required to have continuous enrollment in Medicare Fee for Service for Parts A and B (doctors’ visits and hospitalizations) during follow-up to ensure consistent ascertainment of outcomes, as Medicare Advantage participants do not have data in the CCW file.

Our research is approved by Emory’s IRB (#STUDY00000316) and the Centers for Medicare & Medicaid Services (CMS) under the data use agreement (#RSCH-2020-55733). The Medicare dataset was stored and analyzed in Emory Rollins School secure cluster environment (HPC), with Health Insurance Portability and Accountability Act (HIPAA) compliance.

### Outcome classification

There were two primary outcomes of interest in this study, all-cause dementia and AD. The diagnoses of dementia and AD were identified and recorded as two distinct indicators in the CCW dataset, which incorporated information across the available Medicare claims, including inpatient and outpatient claims, Carrier files (primarily doctor visits), skilled nursing facility, and home health-care claims. The accuracy of this algorithm has been proved in classifying diseases based on several validation studies (20, 21). The ICD codes associated with the two indicators were presented in Table S1. Patients who had dementia diagnoses followed by a later diagnosis of AD were considered to be incident AD cases at the time of their first dementia diagnosis.

### Exposure assessment

15 PM_2.5_ components were explored in this study, including 5 major mass contributors: elemental carbon (EC), organic carbon (OC), nitrate (NO_3_^-^), ammonium (NH_4_^+^), sulfate (SO_4_^2-^), which will be referred to as “major components” afterwards, and 10 trace elements: zinc (Zn) vanadium (V), silicon (Si), lead (Pb), nickel (Ni), potassium (K), iron (Fe), copper (Cu), calcium (Ca), bromine (Br). The annual mean predictions for the 15 PM_2.5_ components was estimated across the contiguous U.S. at a 50 m * 50 m spatial resolution in urban areas and a 1 km * 1 km spatial resolution in non-urban areas from 2000 to 2018 using super-learning and ensemble weighted averaging models. Details about the PM_2.5_ components dataset can be found elsewhere (22-24). Briefly, the training datasets consisted of data from 987 monitoring sites. 166 predictors were used for the super-learning and ensemble weighted-averaging models, including time and geography information, satellite observation data, meteorological data, emitting/surrogate of emission sources and other variables. These approaches achieved excellent model performance, with cross-validated R-square for major components ranging from 0.856 (OM) to 0.957 (SO_4_^2-^) and trace elements ranging from 0.79 (Cu) to 0.88 (Zn). This ensemble model approach has been applied on estimating PM_2.5_ mass concentrations across the contiguous United States before and reached a cross-validated R-square of 0.89 for annual predictions (25). The PM_2.5_ components dataset and PM_2.5_ mass dataset have been widely used in previous epidemiological studies (10, 19, 26).

The gridded predictions for each PM_2.5_ component were then averaged at the ZIP code level with a population weighting approach. Within a ZIP code, the gridded population density value published on NASA SEDAC website were used as weights while calculating the spatial averages of PM_2.5_ components estimates (27). The moving average of up to 5 years prior to each calendar year were calculated by ZIP code. The ZIP code level 5 year moving average of exposures were then assigned to each Medicare beneficiary for each year, based on their residential ZIP code annually.

### Covariates

Demographic characteristics at the individual level, including age, gender, and race, as well as Medicaid insurance status, were sourced from the Medicare denominator file. The age of beneficiaries was categorized into 4 groups: 65-75, 75-85, 85-95, 95+. The analytical models also integrated covariates at the neighborhood level, encompassing ZIP code-based socio-economic status variables such as population density, median household income, the percentage of the Black population, the percentage of the population residing in rental housing or apartments, and the percentage of the population with less than a high school education. Additionally, county-level factors, including behavioral risk factors such as smoking prevalence and mean body mass index, health care capacity variables like the number of hospitals and active medical doctors per 1,000 people, and a geographical region indicator, were incorporated into the models. Comprehensive details and data sources for the covariates could be found in Shi et al. (2021) (7).

### Statistical analysis

Our main analysis assessed the cumulative associations between 15 PM_2.5_ components and two primary outcomes, all-cause dementia and AD using two approaches, weighted quantile sum (WQS) regression models and quantile g-computation (qgcomp) models, with quasi-Poisson link function to account for potential over-dispersion.

The WQS models employ a two-step methodology to analyze the collective impact of exposure to a mixture of pollutants. Firstly, WQS calculates a composite index, representing the pollutant mixture, by deriving a weighted sum of quantiles for each pollutant, with optimal weights estimated by via regression of the studied outcome on the composite index in a training data set. Subsequently, the WQS model estimates the combined association between the pollutant mixture and the outcome through a multivariate regression model, utilizing the WQS term as the exposure metric. This approach provides a comprehensive understanding of the combined effects of multiple pollutants on a specific outcome, incorporating both the composition of the mixture and its impact (28). One of the limitations for WQS model is that it requires directional homogeneity assumption, which means that all the exposures should have same direction associations with the outcome or have null associations.

The qgcomp models were developed as an extension of WQS regression models. Compared to WQS models, the qgcomp models do not require directional homogeneity and can incorporate both positive and negative associations between individual components of the mixture and the outcome. Qgcomp models use a marginal structural model to estimate the effect of the mixture. The qgcomp models can consistently estimate the effects of the exposures when WQS regression might be biased or inconsistent, but may also yield similar estimates with WQS regressions with large samples when its assumptions hold (29). We utilized both WQS and qgcomp models to compare the output from both models and serve as a cross-validation.

Individual level data were aggregated by calendar year and ZIP code in WQS models due to limited computational capacity, while in qgcomp models, individual level covariates (age group, sex, race, Medicaid status) were also used in data aggregation to define a stratum for more precise model estimations. For WQS models, the individual level covariates listed above were converted into the proportion of variable categories in each stratum defined by calendar year and ZIP code.

For WQS, an index was estimated from ranking exposure concentrations in deciles. Then, the dataset was divided by 60:40 into the training dataset and validation dataset. 250 bootstrap samples were assigned for parameter estimation. For qgcomp models. We also converted exposure concentrations into deciles. “qgcomp.noboot” procedure was used to estimate the associations. Both WQS and qgcomp models utilized quasi-Poisson regression, which accounts for any over-dispersion of Poisson assumptions. The observed number of cases in each stratum defined above was the outcome, and the predictors were deciles of the components and covariates. All the neighborhood level covariates were included as linear terms in models unless specified otherwise.

Several sensitivity analyses were also conducted. We fitted single and multi-pollutant stratified Cox proportional hazard models with a generalized estimating equation (GEE) to compare the traditional models with the mixture models. Single pollutant models and multi-pollutant models were fitted using individual level data, stratified by age group, race, sex, and Medicaid status, and controlled for same covariates in WQS and qgcomp models. Besides the model types, we also explored the effect of movers in our study. Since beneficiaries might change their address during the follow-up period, utilizing the moving 5-year average exposures on ZIP code level could introduce exposure misclassification. Thus, we restricted the study population to beneficiaries that never change their address during the follow-up period for mixture models. All analyses were conducted in R software, version 4.2.2. WQS regression models were run with “gWQS” package and qgcomp models were run with “qgcomp” package (30, 31). Statistical significance was determined by two-sided P<0.05.

## Results

### Study Population

Summary statistics of the dementia cohort and AD cohort can be found in Table S1. There were approximately 33.7 million beneficiaries in the dementia cohort and 34.6 million beneficiaries in the AD cohort. About 8.4 million (25.0%) individuals developed dementia and 3.8 million (11.1%) individuals developed AD. Most of the individuals in both cohorts were female (∼57%), White (∼88%), and not eligible for Medicaid insurance (∼90%). Fig 1. shows the county-level incidence of dementia and AD per 100,000 Medicare beneficiaries in United States from 2000 to 2018. A relatively higher incidence of both dementia and AD was observed in the South and Southeast US.

**Figure 1:**
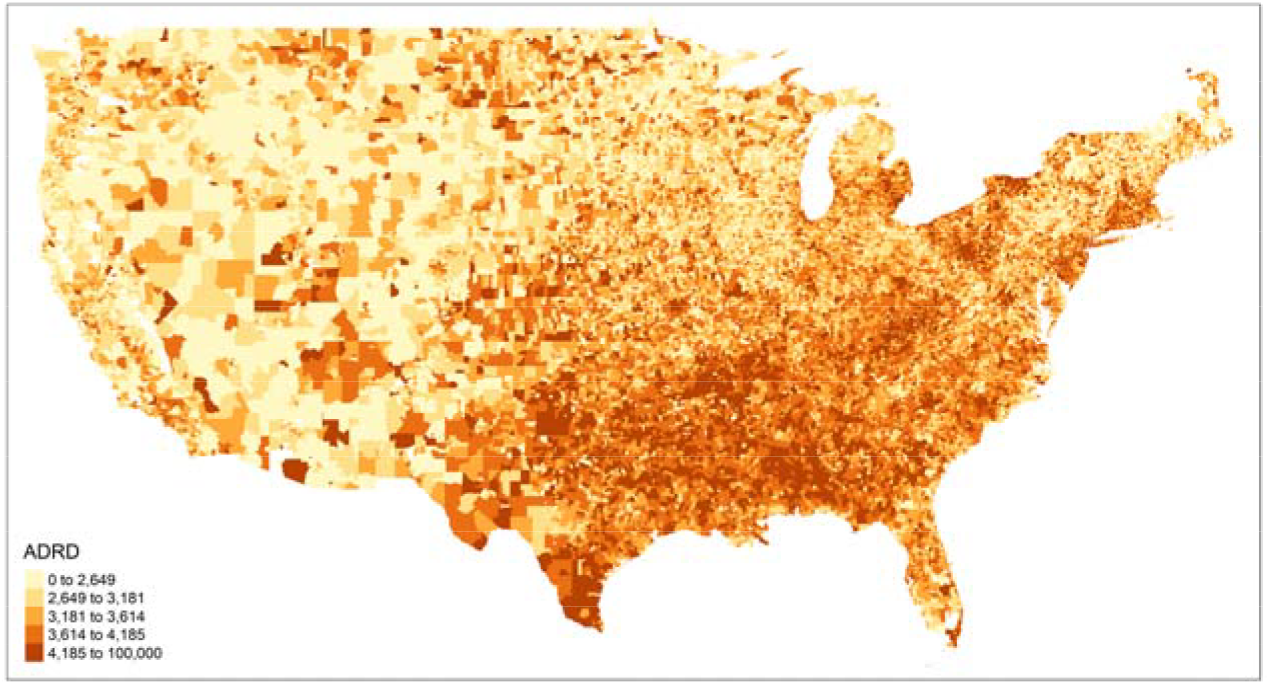
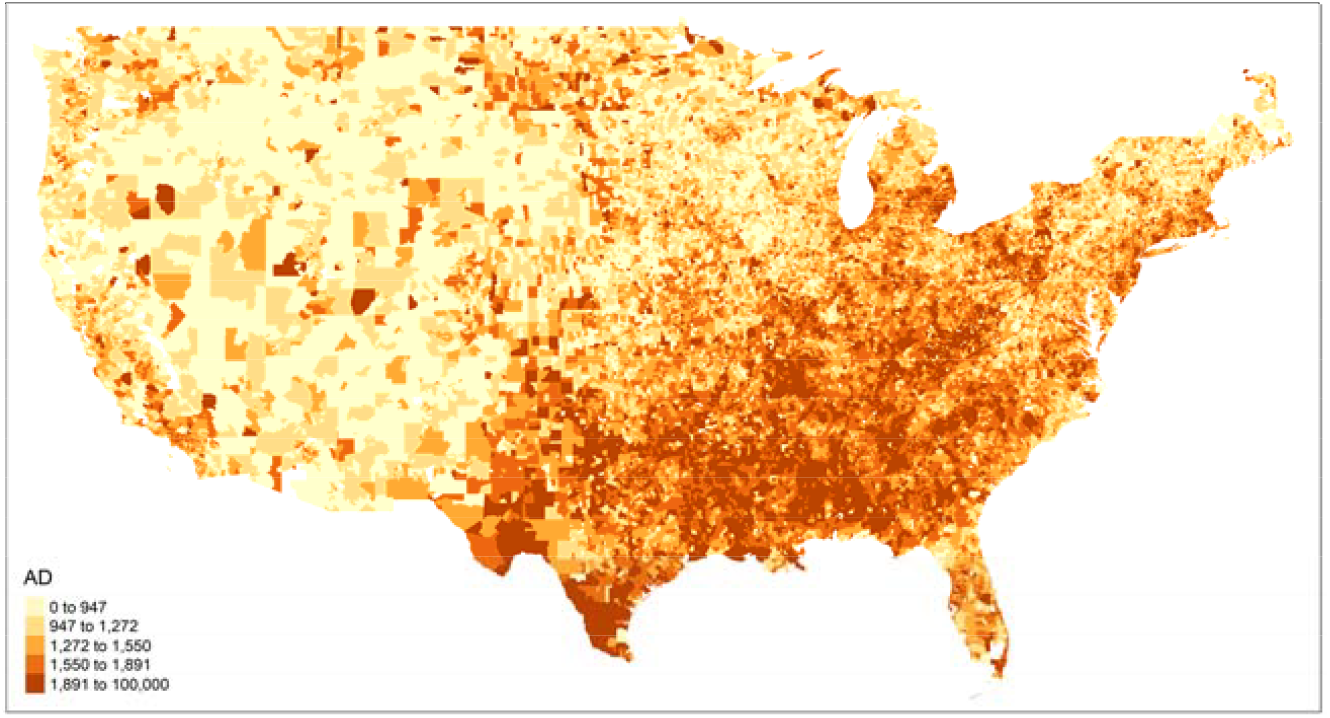
Distribution of Dementia / Alzheimer’s disease per 100000 person-yr by ZIP code across the contiguous U.S.

### Air pollution levels

The annual mean PM_2.5_ mass concentration (available during 2000-2016) was 9.87 μg/m^3^, which was lower than the EPA primary standard level, 12.0 μg/m^3^, with an interquartile range (IQR) of 4.18 μg/m^3^. The mean concentrations of PM_2.5_ components are listed in Table 2. SO_4_^2-^ and OC contributed the most towards the PM_2.5_ mass with mean concentrations of 1.80 μg/m^3^ and 1.56 μg/m^3^ respectively. Among trace elements, Si, Fe, Ca contributed the most, with mean concentrations at 93.01, 49.84, 41.54 pg/m3, respectively. Figure S2 shows the correlation matrix across all 15 PM_2.5_ components. The absolute values of correlation coefficients between PM_2.5_ components ranged from 0.01 to 0.88. Several PM_2.5_ components were highly correlated, such as Cu and EC (correlation coefficient r = 0.80), NH_4_^+^ and SO_4_^2-^ (r = 0.88), Pb and Zn (r = 0.76).

**Table 1:**
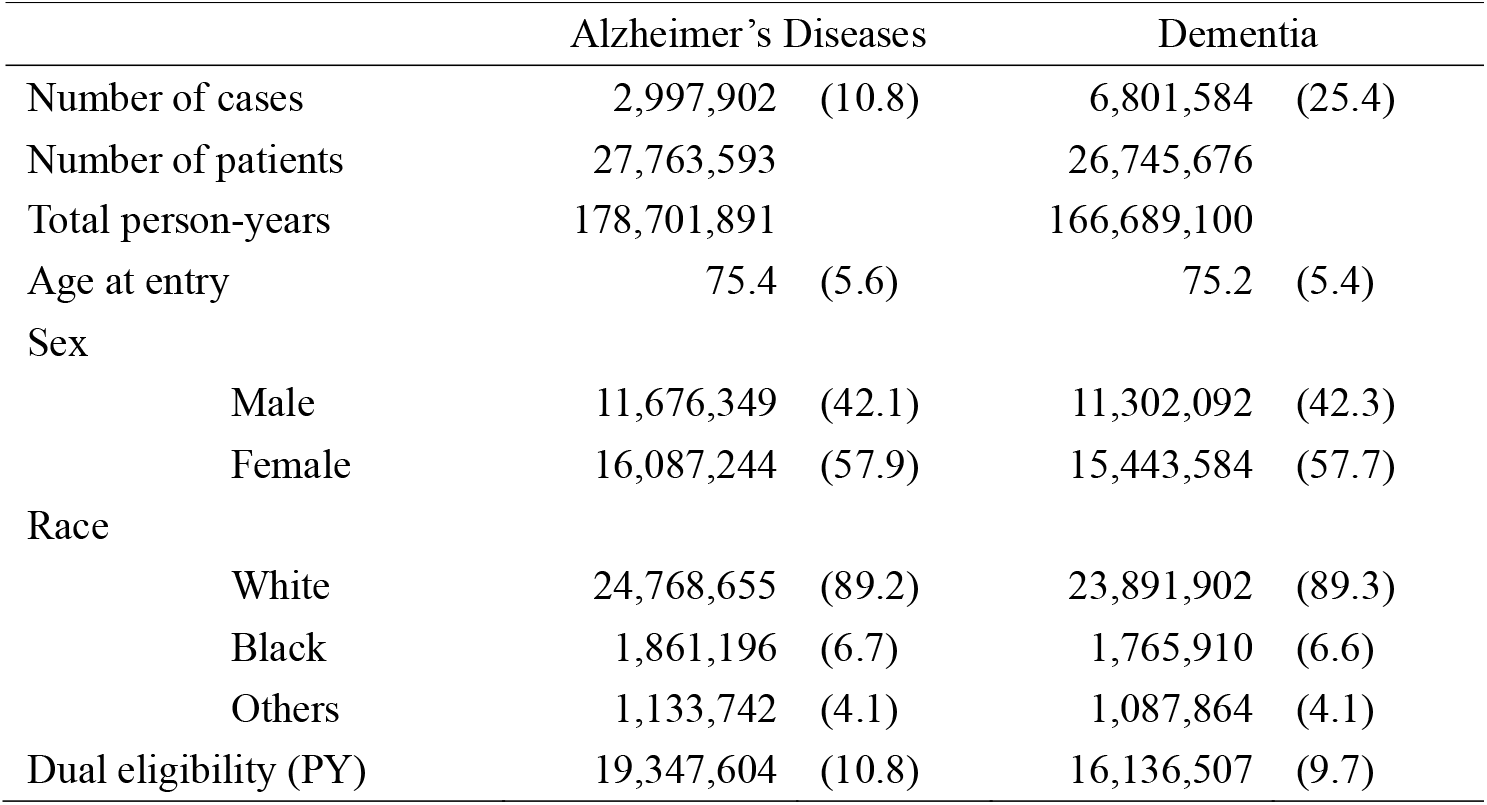
Descriptive statistics of the study population and area-level covariates among the dementia cohort and Alzheimer’s disease (AD) cohort.

|  | Alzheimer's Diseases |  | Dementia |  |
| --- | --- | --- | --- | --- |
| Number of cases | 2,997,902 | (10.8) | 6,801,584 | (25.4) |
| Number of patients | 27,763,593 |  | 26,745,676 |  |
| Total person-years | 178,701,891 |  | 166,689,100 |  |
| Age at entry | 75.4 | (5.6) | 75.2 | (5.4) |
| Sex |  |  |  |  |
| Male | 11,676,349 | (42.1) | 11,302,092 | (42.3) |
| Female | 16,087,244 | (57.9) | 15,443,584 | (57.7) |
| Race |  |  |  |  |
| White | 24,768,655 | (89.2) | 23,891,902 | (89.3) |
| Black | 1,861,196 | (6.7) | 1,765,910 | (6.6) |
| Others | 1,133,742 | (4.1) | 1,087,864 | (4.1) |
| Dual eligibility (PY) | 19,347,604 | (10.8) | 16,136,507 | (9.7) |

**Table 2:** Summary statistics of PM2.5 component concentrations across study period nitrate; NH_4_^+^: ammonium; OC: organic carbon; EC: elemental carbon; Zn: zinc; V or vanadium; K: potassium; Si: silicon; Pb: lead; Ni: nickel; Fe: iron; Cu: copper; Ca: calcium; Br: bromine

| Pollutants | IQR | Mean | SD | 25th | Median | 75th | 95th | Max |
| --- | --- | --- | --- | --- | --- | --- | --- | --- |
| EC (ug/m3) | 0.31 | 0.44 | 0.26 | 0.26 | 0.36 | 0.57 | 0.94 | 2.33 |
| NH<sub>4</sub><sup>+</sup> (ug/m3) | 0.65 | 0.73 | 0.45 | 0.37 | 0.63 | 1.02 | 1.60 | 2.84 |
| NO<sub>3</sub><sup>-</sup> (ug/m3) | 0.71 | 0.92 | 0.56 | 0.50 | 0.77 | 1.22 | 2.00 | 4.83 |
| OC (ug/m3) | 0.75 | 1.56 | 0.64 | 1.12 | 1.44 | 1.87 | 2.75 | 7.14 |
| SO<sub>4</sub><sup>2-</sup> (ug/m3) | 1.48 | 1.80 | 1.07 | 0.98 | 1.53 | 2.45 | 3.99 | 6.47 |
| Br (pg/m3) | 1.18 | 2.22 | 0.83 | 1.63 | 2.21 | 2.81 | 3.59 | 8.09 |
| Ca (pg/m3) | 27.67 | 41.54 | 25.66 | 24.59 | 34.61 | 52.26 | 86.72 | 326.35 |
| Cu (pg/m3) | 1.96 | 1.88 | 1.66 | 0.69 | 1.23 | 2.65 | 5.39 | 17.73 |
| Fe (pg/m3) | 31.98 | 49.84 | 28.43 | 30.59 | 42.39 | 62.57 | 106.99 | 241.28 |
| K (pg/m3) | 18.20 | 51.62 | 15.42 | 41.94 | 50.65 | 60.15 | 76.77 | 232.27 |
| Ni (pg/m3) | 0.53 | 0.45 | 0.49 | 0.11 | 0.33 | 0.63 | 1.26 | 6.23 |
| Pb (pg/m3) | 1.45 | 2.05 | 1.29 | 1.16 | 1.62 | 2.61 | 4.82 | 9.72 |
| Si (pg/m3) | 63.28 | 93.01 | 53.02 | 55.55 | 80.30 | 118.83 | 187.00 | 581.00 |
| V (pg/m3) | 0.59 | 0.61 | 0.73 | 0.17 | 0.35 | 0.76 | 2.15 | 9.33 |
| Zn (pg/m3) | 4.30 | 6.78 | 3.89 | 4.22 | 5.92 | 8.51 | 13.87 | 46.59 |
| Total PM<sub>2.5</sub> mass (ug/m3) | 4.18 | 9.87 | 3.17 | 7.76 | 9.76 | 11.94 | 15.10 | 30.92 |

Note: PM<sub>2.5</sub> mass concentrations are available from 2000 to 2016. Abbreviations: IQR: interquartile range. SD: Standard deviation. 25<sup>th</sup>: 25<sup>th</sup> percentile. 75<sup>th</sup>: 75<sup>th</sup> percentile. 95<sup>th</sup>: 95<sup>th</sup> percentile. SO<sub>4</sub><sup>2-</sup>: sulfate; NO<sub>3</sub><sup>-</sup>:

### Qgcomp and WQS models on 15 PM_2.5_ components

The cumulative association between PM_2.5_ components and dementia/AD were summarized in Table 3. From the qgcomp models, the rate ratio for dementia was 1.027 (95% CI: 1.025-1.028, per 1 decile increase in all PM_2.5_ components), while AD showed stronger associations with a rate ratio of 1.041 (95% CI: 1.039-1.042). Similar but slightly smaller estimates were obtained from WQS models, with the rate ratio for dementia at 1.029 (95% CI: 1.028, 1.030) and AD at 1.048 (95% CI: 1.047, 1.049) per 1 decile increase in all PM_2.5_ components.

**Table 3:** Cumulative associations between PM_2.5_ components and Alzheimer’s diseases / dementia in rate ratios estimated from both WQS and qgcomp models.

|  | WQS | qqcomp |
| --- | --- | --- |
| Dementia | 1.029 (1.028, 1.030) | 1.027 (1.025, 1.028) |
| Alzheimer's diseases | 1.048 (1.047, 1.049) | 1.041 (1.039, 1.042) |

Note: The rate ratios are estimated with 1 decile increase in all $\text{PM}_{2.5}$ components.

The weights of each PM_2.5_ component contributing to the associations for dementia and AD in qgcomp and WQS models were shown in Figure 2 and 3, respectively. Cu, SO_4_^2-^, and OC were among the highest weights for both dementia and AD in both qgcomp and WQS models. In addition, qgcomp model identified Fe for AD and Zn for dementia with relatively large positive weights; WQS model identified Ni for dementia for relatively large weights. The weights of other components were very small and negligible.

**Figure 2:**
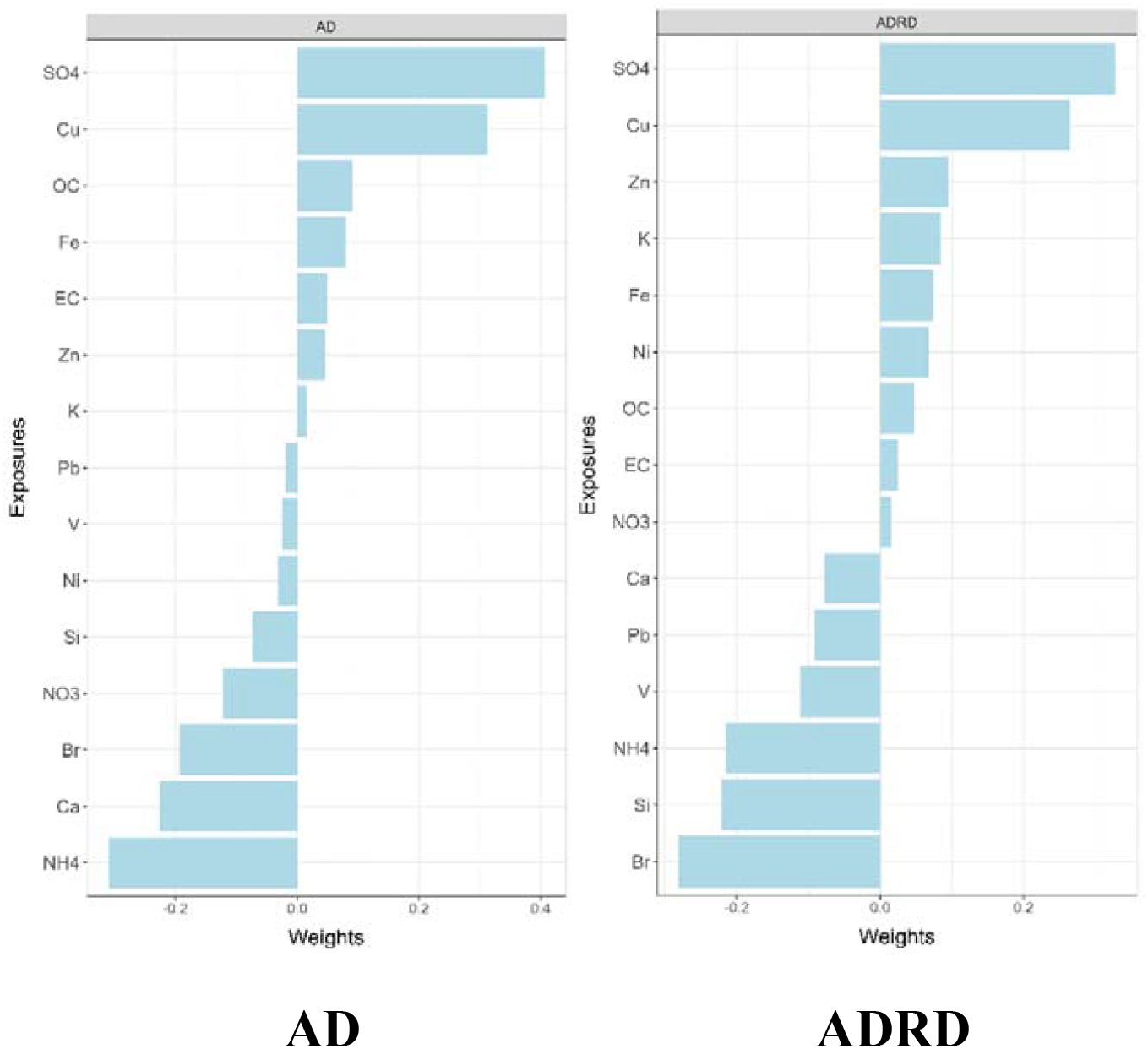
The weights assigned to each PM_2.5_ component from qgcomp models.

**Figure 3:**
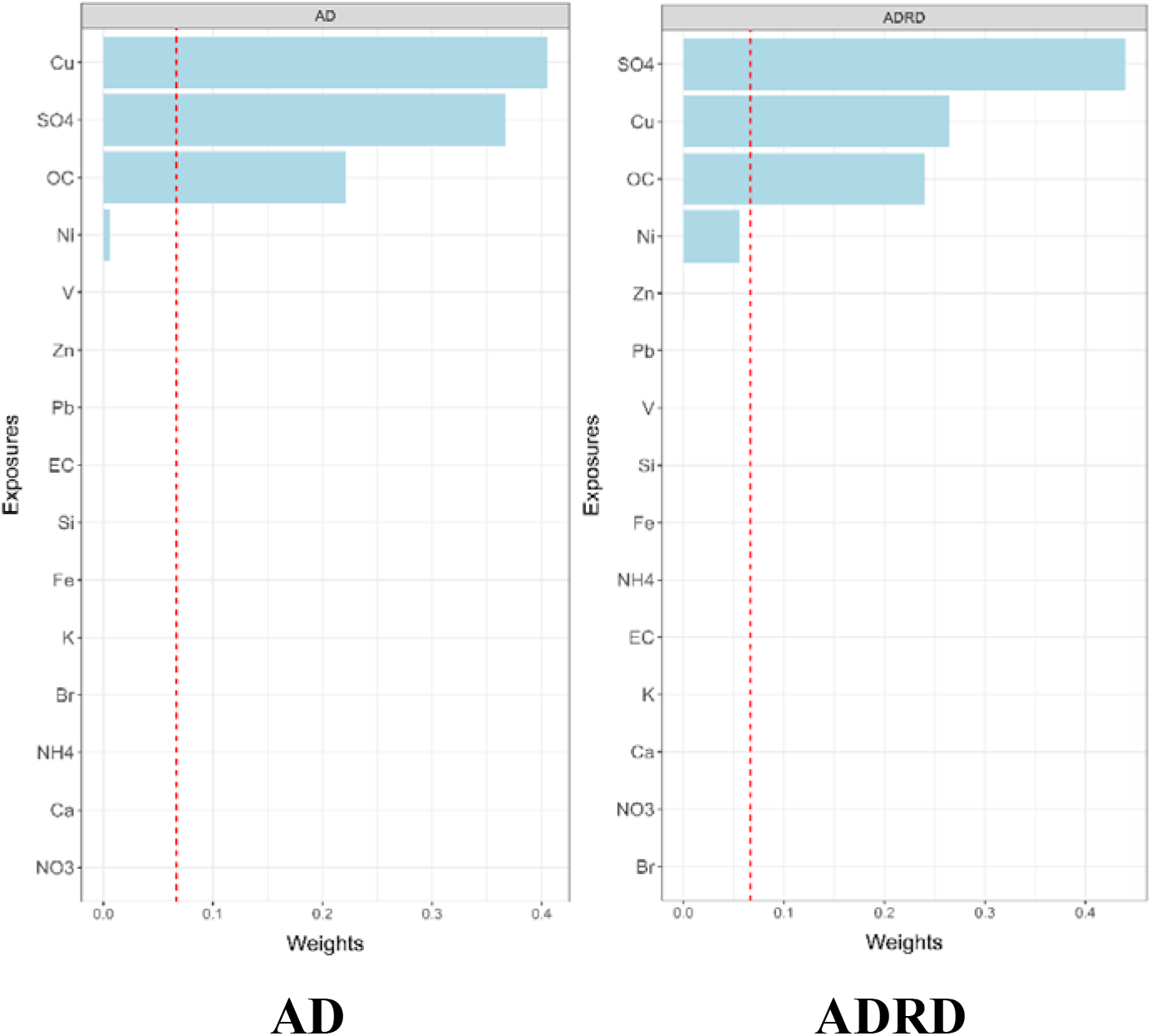
The weights assigned to each PM_2.5_ component from WQS models. Red dashed line marks the value of 1/15, which is the threshold for identifying significant contributor among all PM_2.5_ components.

### Total PM_2.5_ mass and single component cox model

The total PM_2.5_ mass and single PM_2.5_ component results are presented in Figure S4 and Table S3. PM_2.5_ components with highest weights in mixture models also showed relatively strong association in single pollutant models. The hazard ratios of dementia with per IQR increase in PM_2.5_ components were 1.057 (95% CI: 1.052, 1.062) for Cu, 1.086 (95% CI: 1.081, 1.091) for SO_4_^2-^, and 1.042 (95% CI: 1.037, 1.047) for OC. Similar but stronger associations were found for AD with HRs of 1.073 (95% CI: 1.067, 1.080) for Cu, 1.099 (95% CI: 1.091, 1.107) for SO_4_^2-^, 1.066 (95% CI: 1.059, 1.074) for OC.

For Zn, we observed higher HR = 1.052 (95% CI: 1.048, 1.055) for dementia but smaller HR for AD (1.043, 95% CI: 1.038, 1.048), which was similar to the results in mixture models that Zn had relatively large weight for dementia but much smaller for AD. For Fe, we observed HR = 1.033 (95% CI: 1.028, 1.038) for dementia, and 1.051 (95% CI: 1.045, 1.057) for AD, which also matched with the higher weights for AD identified in qgcomp models.

Additionally, the single pollutant models identified EC (HR = 1.065, 95% CI: 1.060, 1.071), NH_4_^+^ (HR = 1.060, 95% CI: 1.055, 1.065), Br (HR = 1.042, 95% CI: 1.037, 1.047) with relatively strong association with dementia. Similar associations were observed for AD and such associations were stronger for EC and Br, but slightly weaker for NH_4_^+^.

### Sensitivity analysis

The sensitivity analyses results were presented in Table s6. When restricting the study population to non-movers, the estimated rate ratio per deciles was 1.022 (95% CI: 1.020, 1.023) for dementia and 1.036 (95% CI: 1.034, 1.038) for AD in qgcomp models. Both estimates were slightly smaller than those from the full study population. The weights under non-mover scenario were presented in Figure s6. Compared to the main results, SO_4_^2-^ still has the largest weights, while the weight for Cu decreased in qgcomp models for dementia. Additionally, the weights of Fe increased in qgcomp models, and WQS models identified Fe as significant contributor for dementia.

When changing the exposure window from 5 years to 3 years, the estimated rate ratio almost did not change. The weights under 3-year exposure window were presented in figure s7. For weights in qgcomp models, SO_4_^2-^ and Cu remained the highest contributors, while the weights of other components varied slightly. The weights of OC decreased but still positively contributed to the total effect. The weights in WQS models almost did not change.

## Discussion

In this study, we found positive cumulative associations between the PM_2.5_ components and both dementia and AD in mixture analyses. Such positive associations persist across different modeling approaches and specifications. The associations for AD were generally stronger than those for dementia. In both WQS and qgcomp models, SO_4_^2-^, Cu and OC were identified as the main contributors to both dementia and Alzheimer’s disease. Those three components also showed positive associations in both single and multi-pollutant Cox models and remained as important contributors under most of the sensitivity analysis scenarios.

The positive cumulative associations between PM_2.5_ components and both AD and dementia are consistent with findings from our previous study on the Medicare beneficiaries, which utilized PM_2.5_ mass as the primary exposure, and many other epidemiological studies (7, 32-34). The relatively weaker associations for dementia compared to AD might result from the broader spectrum of different mental disorders of dementia. Some of the non-AD disorders under dementia might have weaker association with PM_2.5_ exposures (35). The magnitude of the cumulative associations was much higher when converted to per IQR increase in all components compared to that for total PM_2.5_ mass. Such differences could be due to the temporal and spatial variations in the composition of PM_2.5_. When the PM_2.5_ mass level changes, different components might not change in the same magnitude, potentially resulting in less overall health effects.

Cu, SO_4_^2-^ and OC were characterized as the most important contributing factors to incidence of AD and dementia among all the 15 PM_2.5_ components. Such results were also supported by the single pollutant cox models. It is well-established that PM_2.5_ is small enough to be absorbed into blood circulation through inhalation, and can penetrate the blood-brain barrier (BBB) (36), which indicates that the components in the PM_2.5_ mixture could potentially participate in the metabolism of the nervous system. SO_4_^2-^ and organic carbon are two major PM_2.5_ components that contribute to large fractions in PM_2.5_ mass. The primary sources of SO_4_^2-^ include fossil fuel combustion, industrial process and the fertilizers used in agriculture (37, 38). While there are not many studies reporting the effect of SO_4_^2-^ exposure on nervous system, SO_4_^2-^ could serve as a proxy for fossil fuel combustion or industrial related emission sources. Such sources produce volatile organic compounds (VOC) which could cause oxidative stress and neuroinflammation (39), and further contribute to AD and dementia.

In terms of OC, the exact compound and the emission sources were more complex. OC could be categorized into primary OC, which is mainly produced by vehicle emissions, industrial processes and wildfires, etc., and secondary OC, which is formed through chemical reactions involving VOCs (40). The neurotoxicity of different compounds in OC could differ a lot. For example, PAHs and PCBs in OC are highly toxic (41, 42). More research on the health effects of OC on nervous systems are warranted.

Cu along with Zn and Fe that were identified by qgcomp models are trace elements in PM_2.5_ mixtures. Although trace elements only account for a small part in PM_2.5_ mixtures, they could still have important health impacts. Previous research has shown that Cu could play an important role in the development and progression of neurodegenerative diseases (43). Excessive exposure to Cu could result in the dysregulation of Cu homeostasis in brain tissues (44). Cu overload can cause oxidative stress which could further cause damage to brain cells (45). Fe is an essential cofactor for many proteins in brain, however, various studies have shown that the abnormally high concentrations of Fe in some brain tissues were associated with neurodegenerative disorders (46, 47). Elevated Fe levels in brain could contribute to β-amyloid (Aβ) dysfunction, formation of plaque, the hyperphosphorylation of tau protein, and neuronal cell death which are the classical features in the pathology of AD (48-50). Zn is also an important element in neurons. Disruptions to Zn homeostasis has been associated with multiple nervous system disorders such as AD and Parkinson’s disease (51). Elevated Zn levels in brain tissue could also lead to the deposition of Aβ and overly phosphorylated tau protein, which contributed to AD (52, 53). Considering the plausible adverse health effects of Cu, Zn and Fe on AD and dementia, policies targeting the related emission sources are vital. For example, non-tailpipe emissions from brake and tire wear are significant contributors to the metal components in particulate matter, including Cu, Zn and Fe (54, 55). With the implementation of mobile source regulations and the growing adoption of electric vehicles (EVs), significant reductions in tailpipe emissions have been achieved, and the relative contribution of non-tailpipe emissions has been increasing (56). Targeting the non-tailpipe emissions, EPA has introduced the Copper-free Brake Initiative since 2015 (57), and we call for more effort in the reduction of non-tailpipe emissions.

While the epidemiological association between PM_2.5_ mass and dementia has been extensively studied, there are not many studies looking into the component-specific associations, especially with the mixture analysis techniques. Our previous study revealed the potential effects of SO_4_^2-^ and black carbon (BC) on dementia with traditional cox models (19). More studies that focus on the PM_2.5_ components or PM_2.5_ sources are warranted to better elucidate the health effects of different components in PM_2.5_ mixtures and contribute to targeted policies on PM_2.5_ sources.

There are several strengths in our study. First, our study utilized nationwide, population-based open cohort constructed from the Medicare database to explore the potential health effects of PM_2.5_ mixtures on dementia and AD. The large sample size provided us with strong power to identify the potential health effects. Second, we used mixture analysis techniques, qgcomp and WQS, to avoid the biases derived from collinearity in multi-pollutant models and address the issue of confounding from other PM_2.5_ components in single pollutant models. By comparing the results from mixture analysis methods and traditional methods, our study could provide a more solid insight of the associations between PM_2.5_ components and dementia. Third, our study used comprehensive Medicare claims with physician visits included, which could better capture the earlier diagnosed cases. Compared to other studies that used hospitalization records, our study could better identify incident cases.

There are also several limitations in our study. First, we used predicted PM_2.5_ components concentrations from machine learning models. Though the model performance was excellent, potential prediction errors still exists. Meanwhile, the linkage of exposure to Medicare beneficiaries was conducted on ZIP code level since we only had the residential ZIP code of each beneficiary enrolled in Medicare. Although we have used population-weighted exposures within each ZIP code, some exposure misclassification is inevitable. Second, the qgcomp and WQS models were quantile-based models, and the overall estimates are based on each PM_2.5_ component increase by the same quantile. However, in real-life situations, different PM_2.5_ components rarely change in similar ways, which makes the overall estimates harder to interpret. Meanwhile, we did not account for the interactions among different components nor non-linear functions of the components. Finally, we did not account for the potential outcome misclassification in this study. Since the progression of dementia and AD is a long process, the identification of incident cases was still challenging although we have used comprehensive claims and a 5-year clean period. Misdiagnoses of AD and other dementia has been reported among Medicare beneficiaries before (21). Potential misclassification of AD outcome in Medicare was investigated via two different methods in our previous study on the health effects of PM_2.5_ mass on AD and dementia, and revealed that such outcome misclassification would like to lead to some modest under-estimation of the true hazard ratios (7)..

## Conclusion

Our study revealed a strong positive association between the PM_2.5_ components mixture with both dementia and Alzheimer’s diseases. Cu, SO_4_^2-^ and OC were identified as potentially more important contributors among the 15 PM_2.5_ components. Our findings suggested that policies that target the reduction of ambient PM_2.5_ concentrations should continue to be implemented to reduce the burden of dementia and Alzheimer’s diseases in US. Our findings suggest that reducing PM_2.5_ emissions from traffic and fossil fuel combustion could help mitigate the growing burden of dementia and Alzheimer’s disease.

## Supporting information

Supplemental tables and plots

## Data Availability

The PM2.5 data that support the findings of this study are publicly available from
https://doi.org/10.7927/0rvr-4538, NO2 and O3 data are available from https://doi.org/
10.6084/m9.figshare.16834390. Behavioral risk factors are publicly available from https://
www.cdc.gov/brfss/annual_data/annual_data.html; SES data are publicly available from https://
www.census.gov/data/datasets/2000/dec/summary-file-3.html, https://www.census.gov/
data/datasets/2010/dec/summary-file-1.html, and https://www.census.gov/data/
developers/data-sets/acs-1year.html. Health-care capacity data are available from https://
data.hrsa.gov/topics/health-workforce/ahrf. The rules governing the main Medicare dataset
used in this study prohibit any sharing of the health datasets being used here, restricted by for our Data Use Agreement with the US Centers for Medicare & Medicaid
Services. Academic and non-profit researchers who are interested in using
Medicare data should contact the US Centers for Medicare & Medicaid Services directly
to obtain their own datasets upon completion of a Data Use Agreement.

## Acknowledgements

This study was supported by the National Institute on Aging (NIA/NIH R01 AG074357).

