## Supplemental tables and plots for "The role of the components of PM_2.5_ in the incidence of Alzheimer’s disease and related disorders"

Figure S1: ZIP code level mean concentrations (ug/m^3^) of 15 PM_2.5_ components across 2000-2018

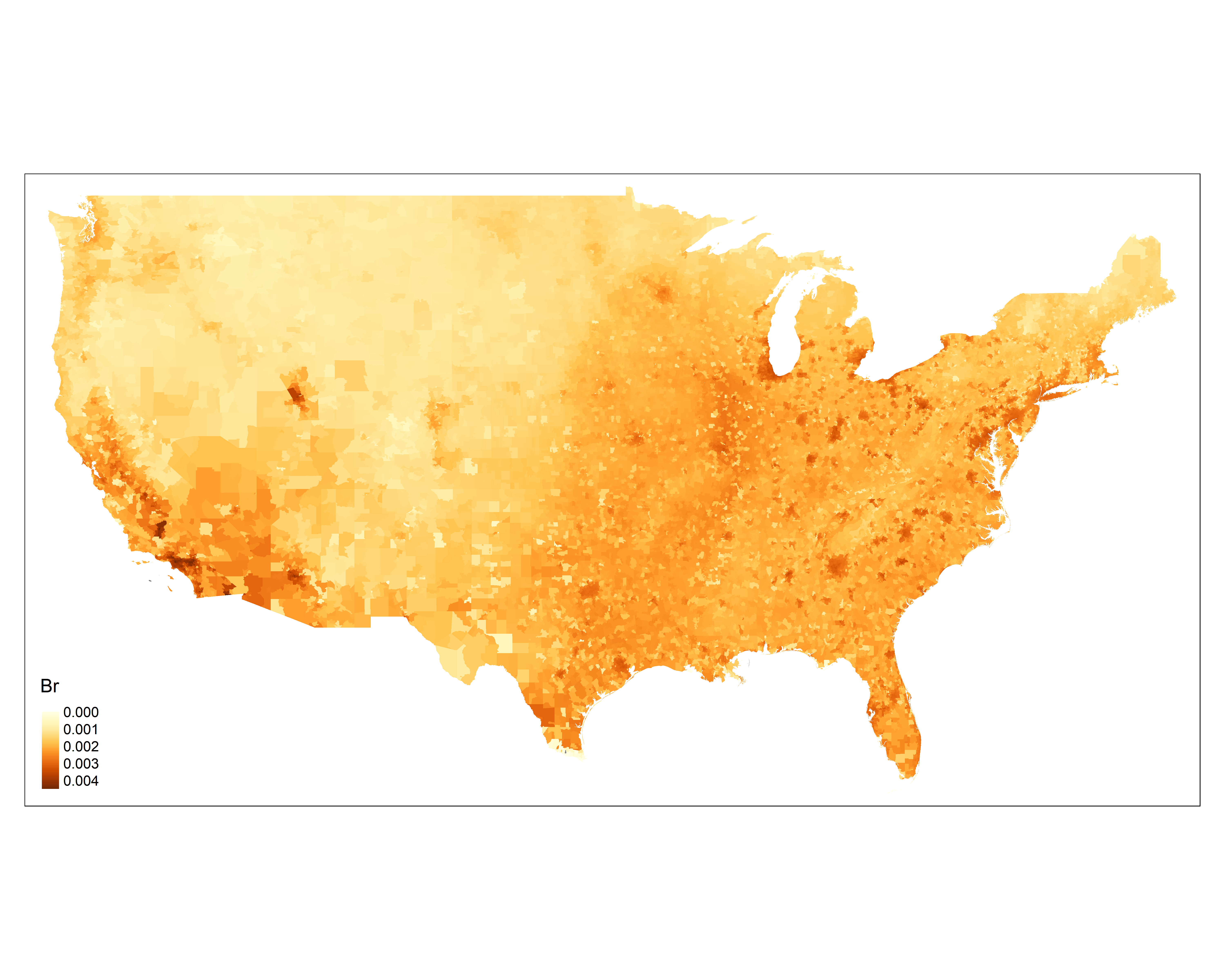

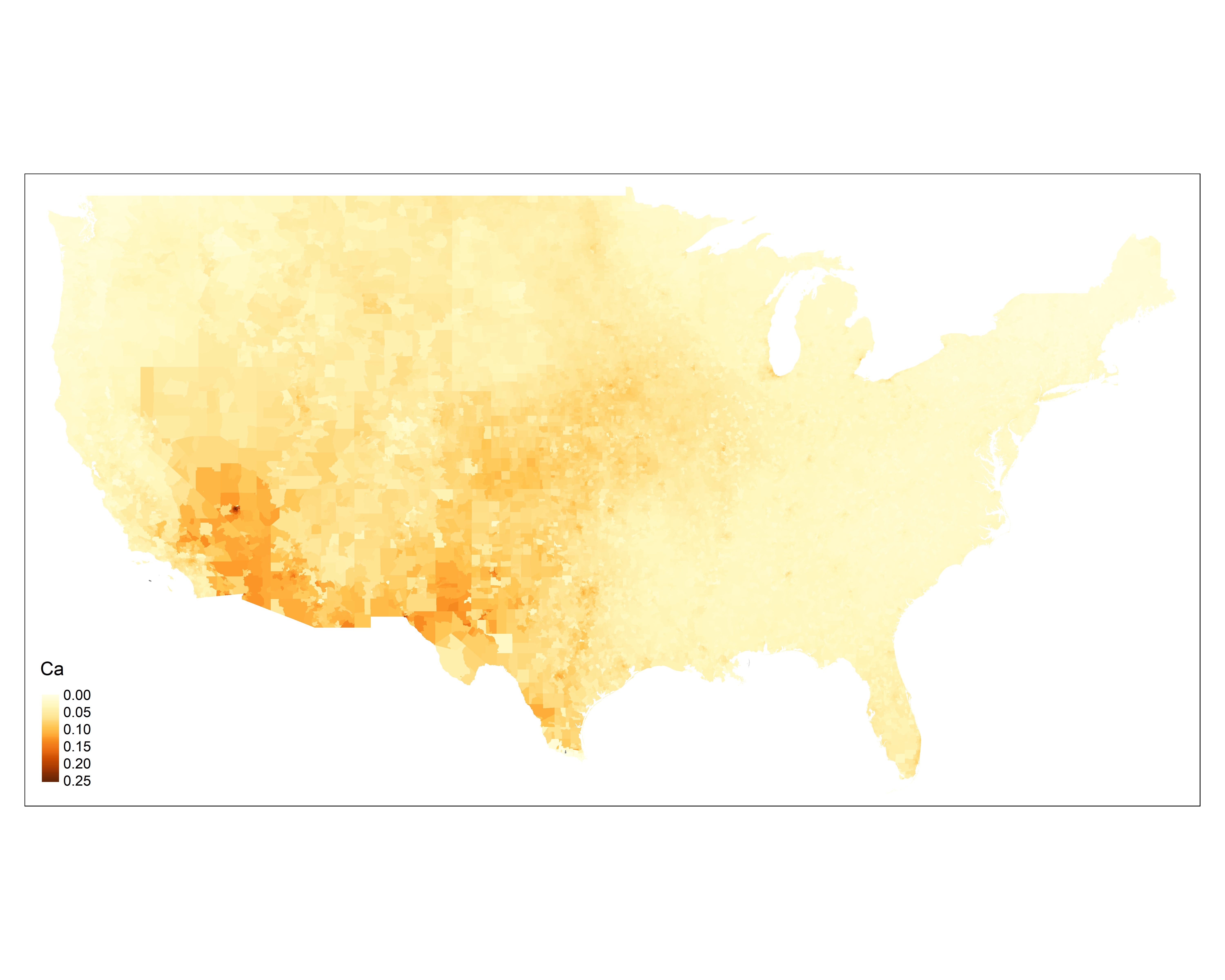

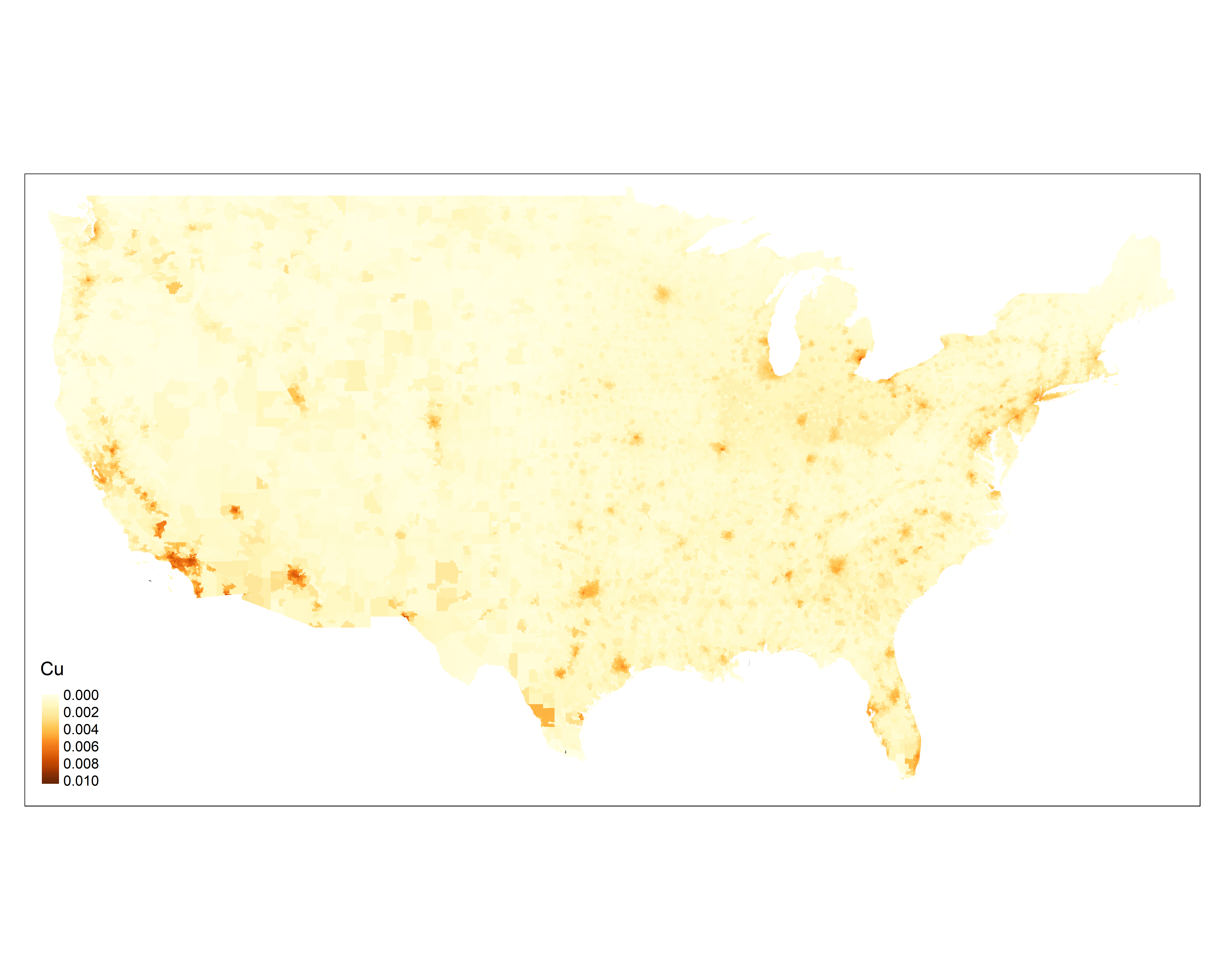

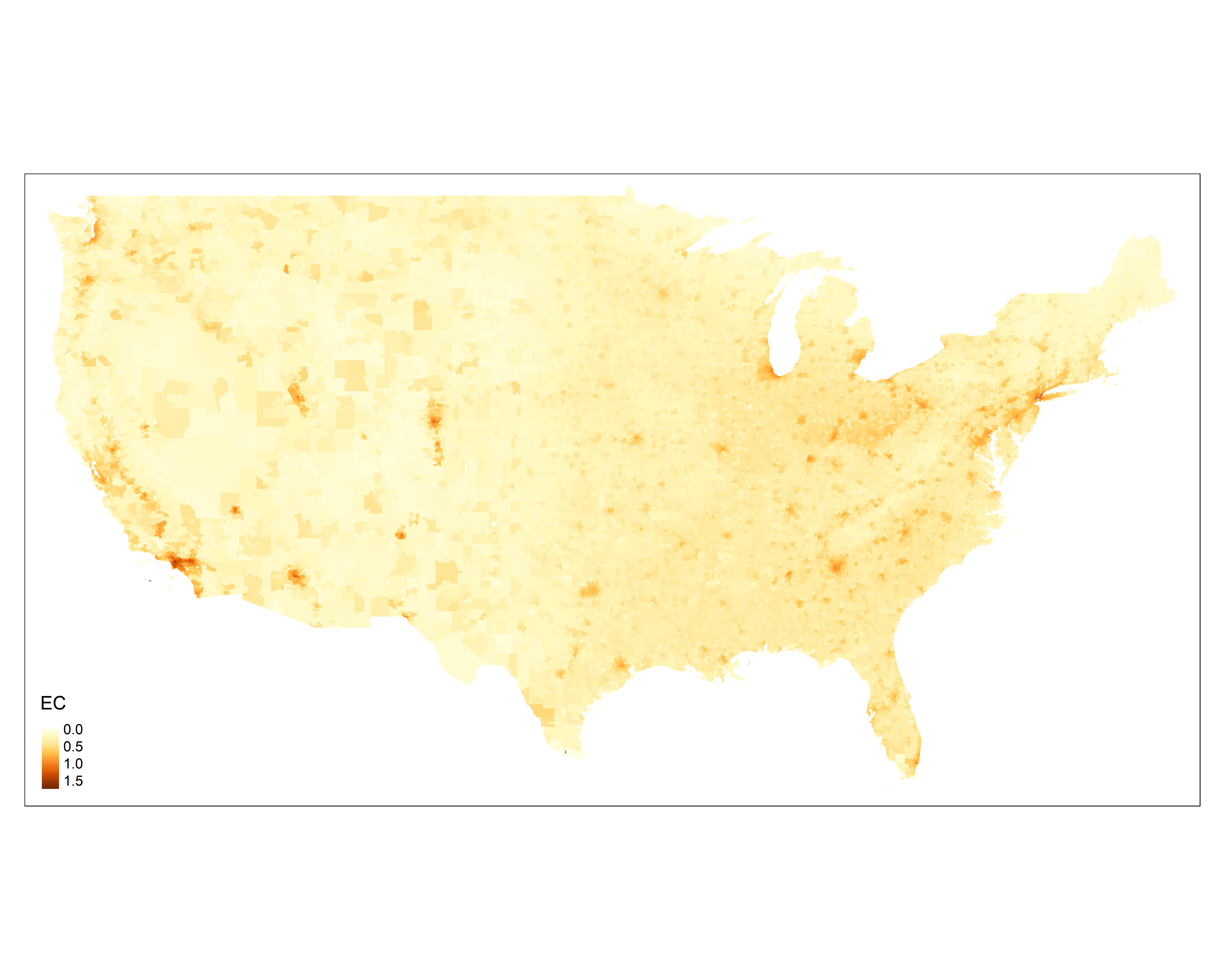

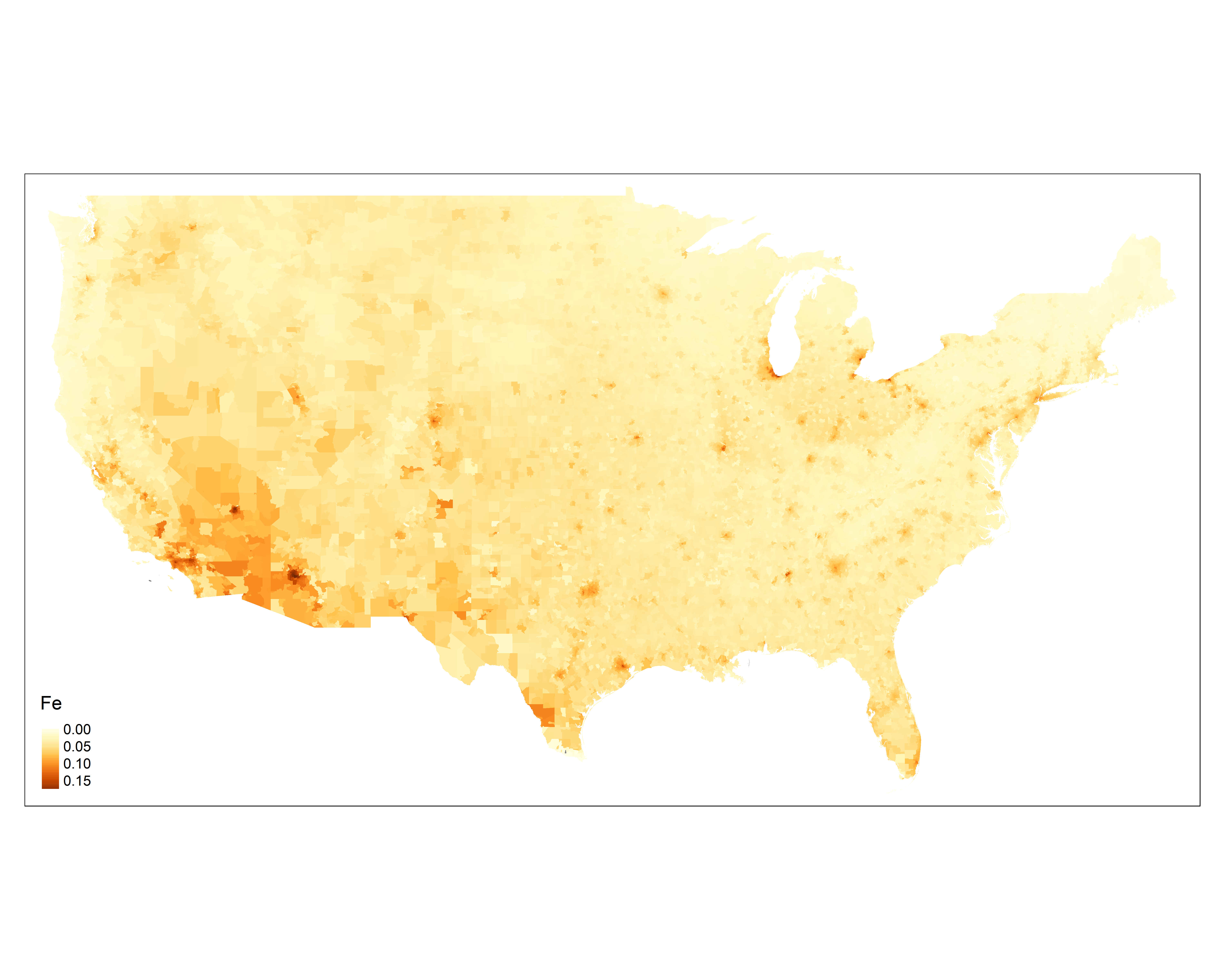

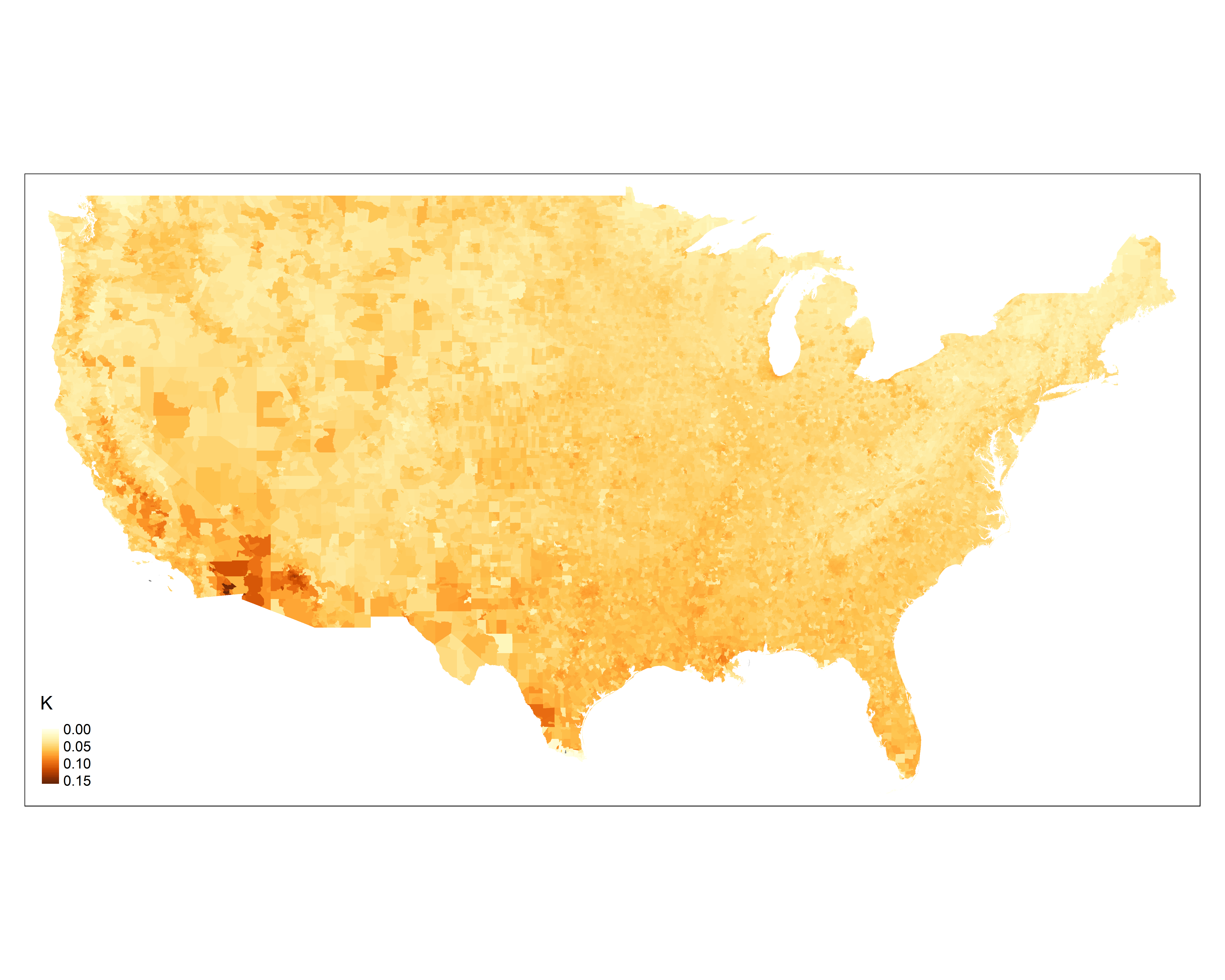

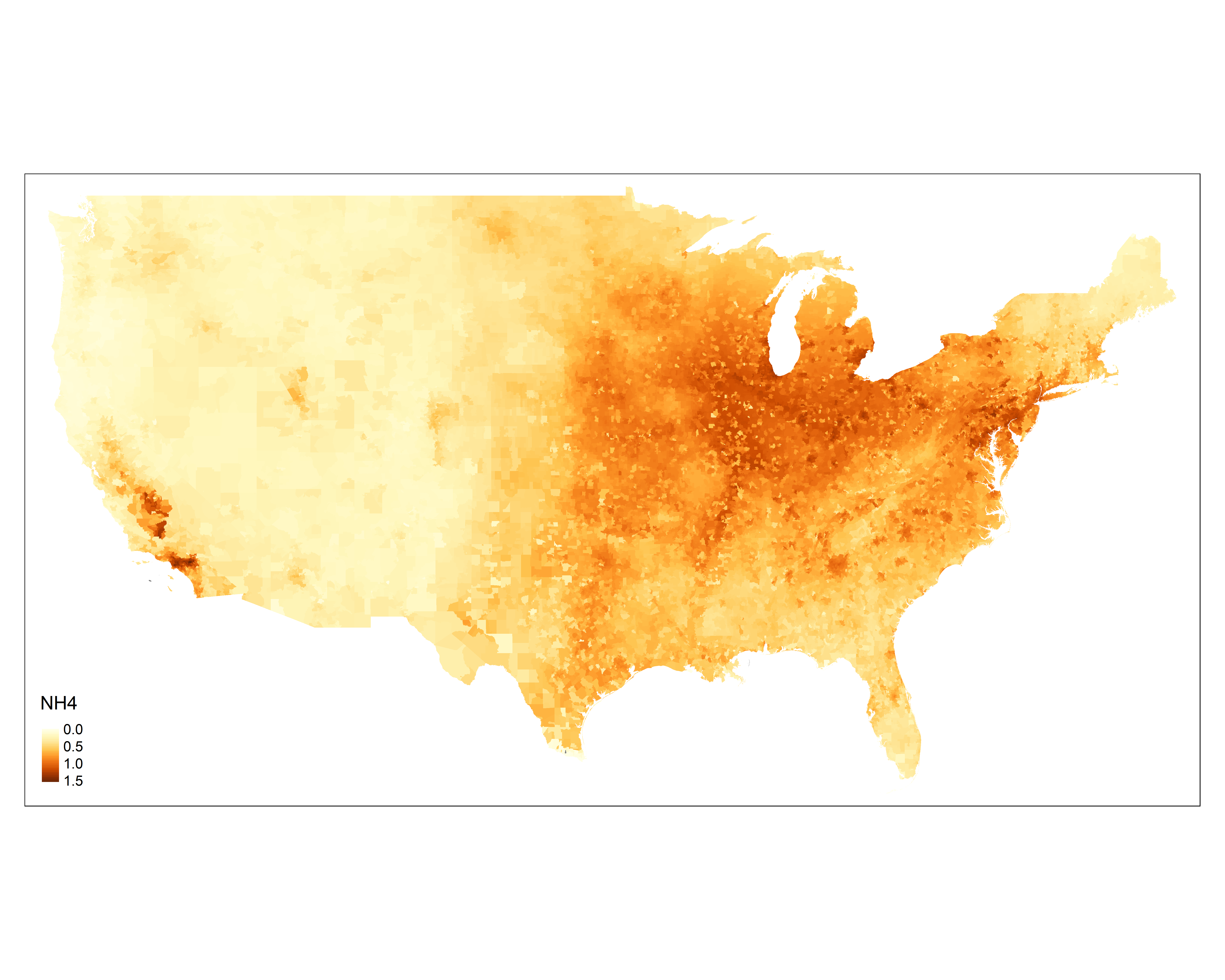

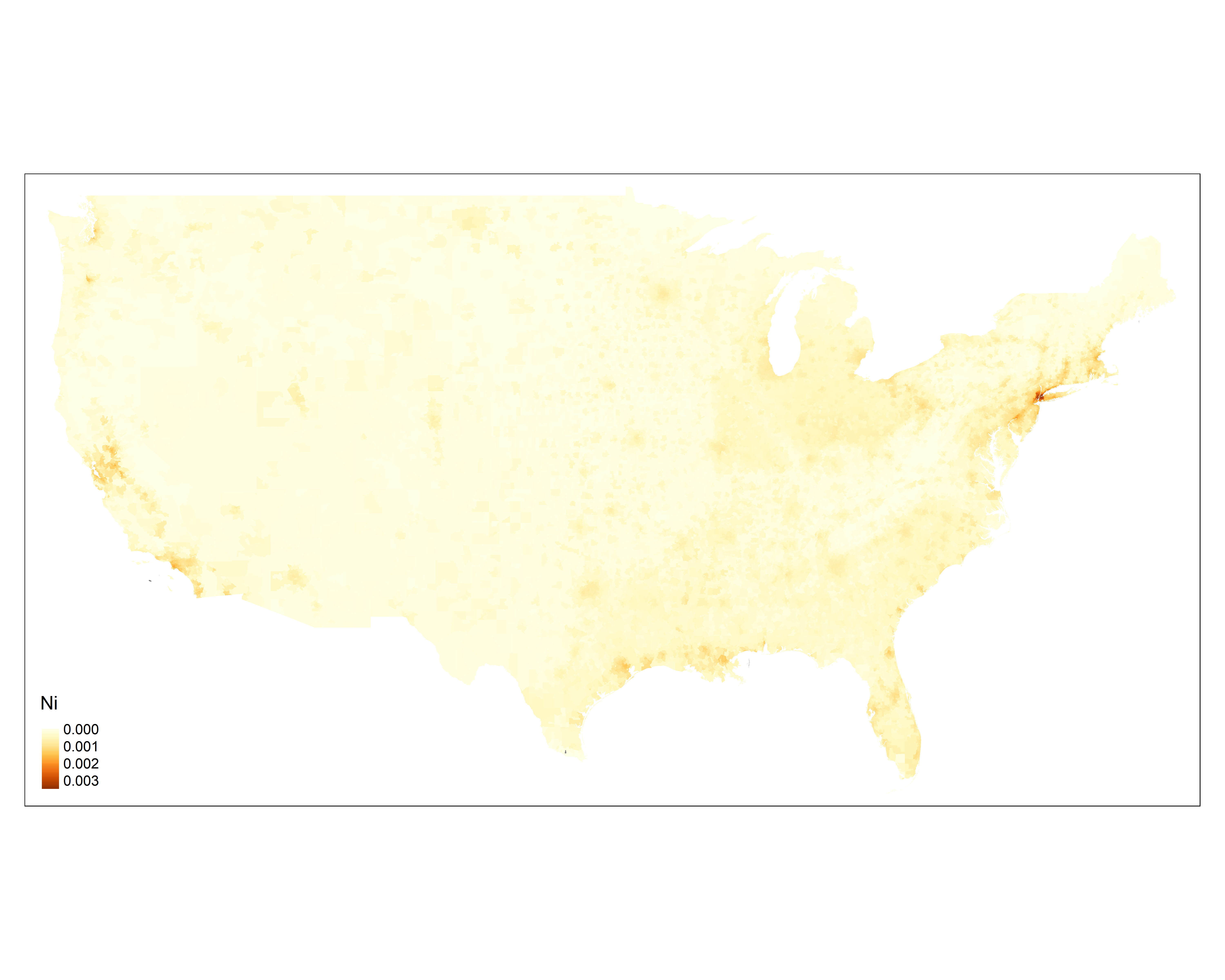

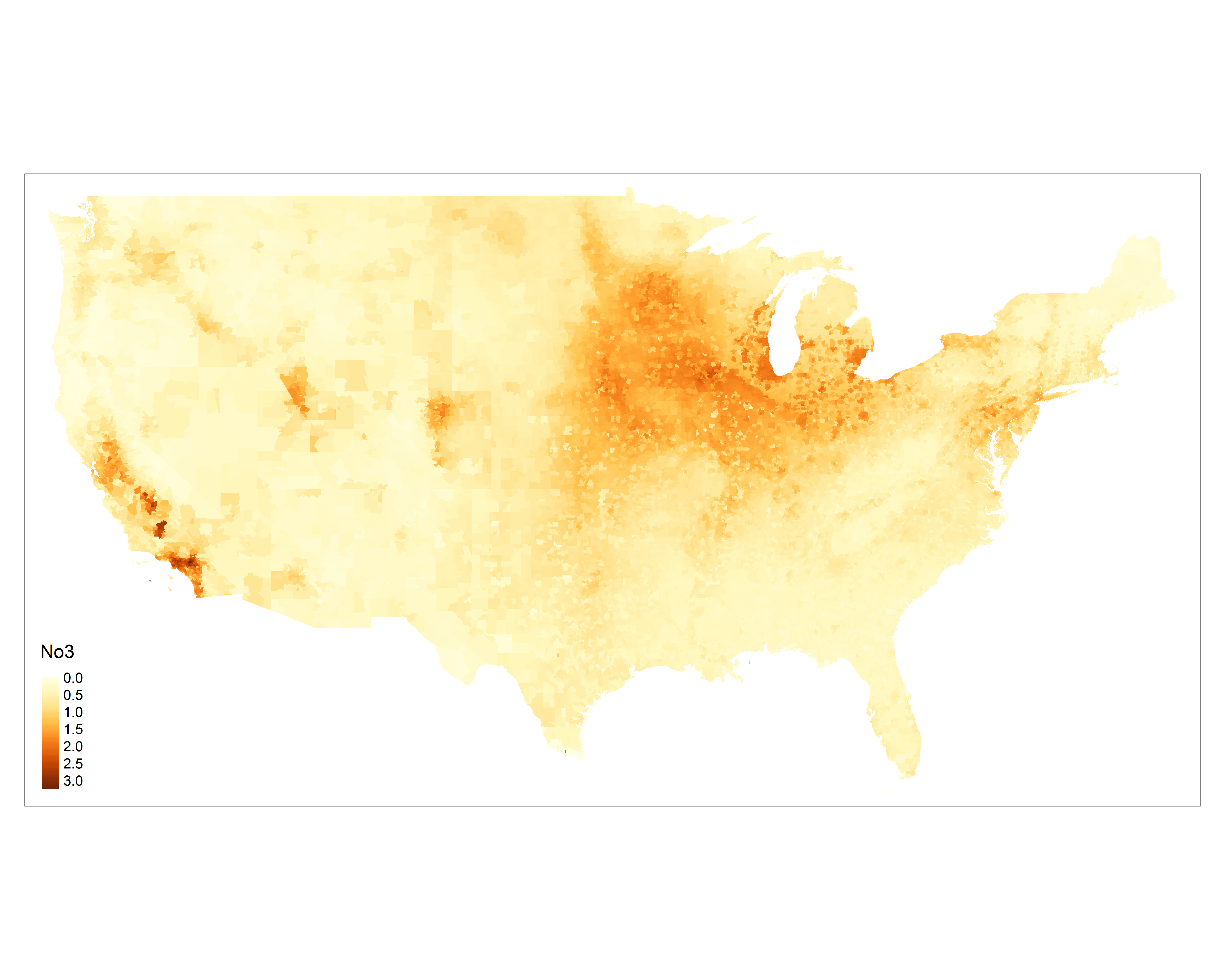

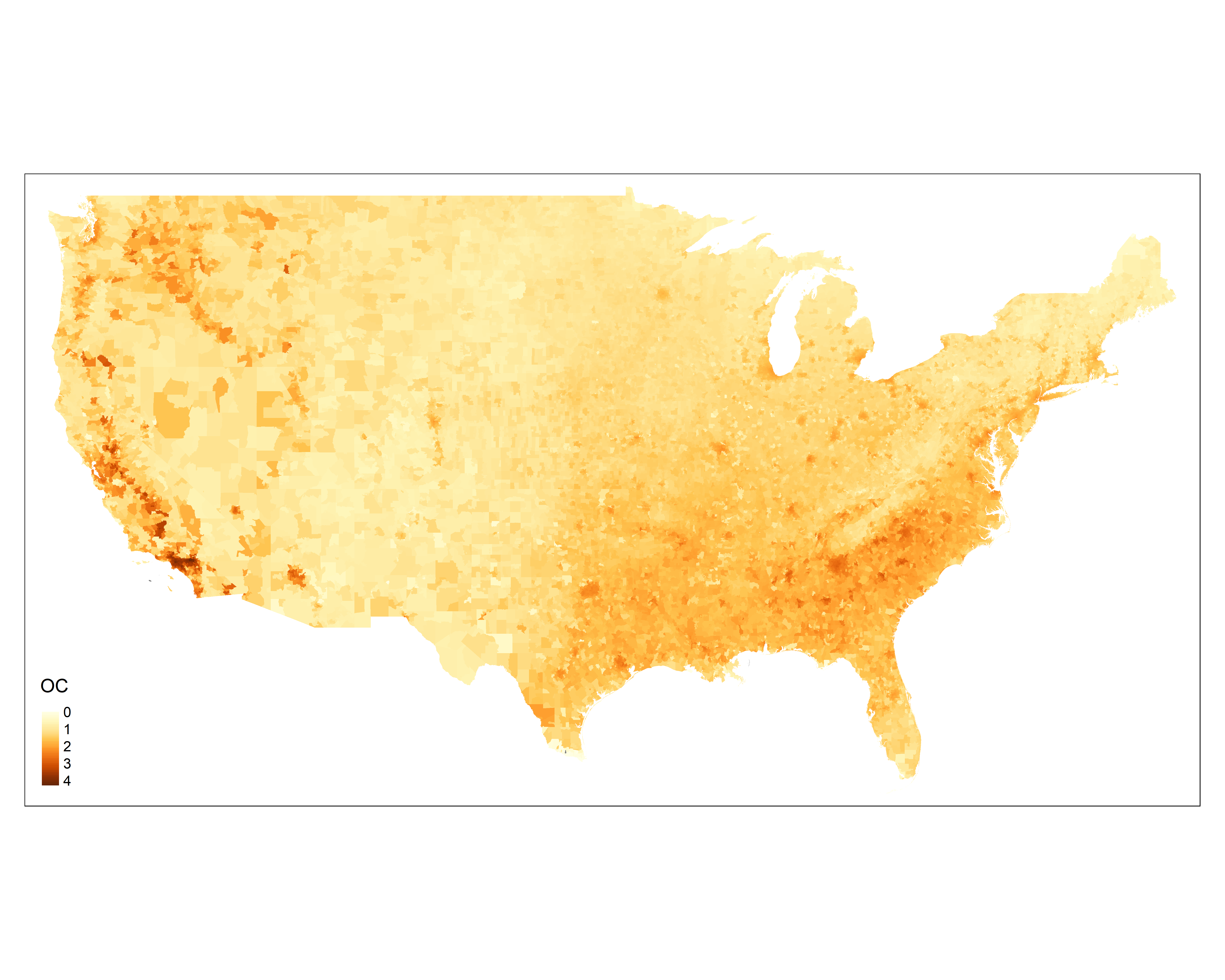

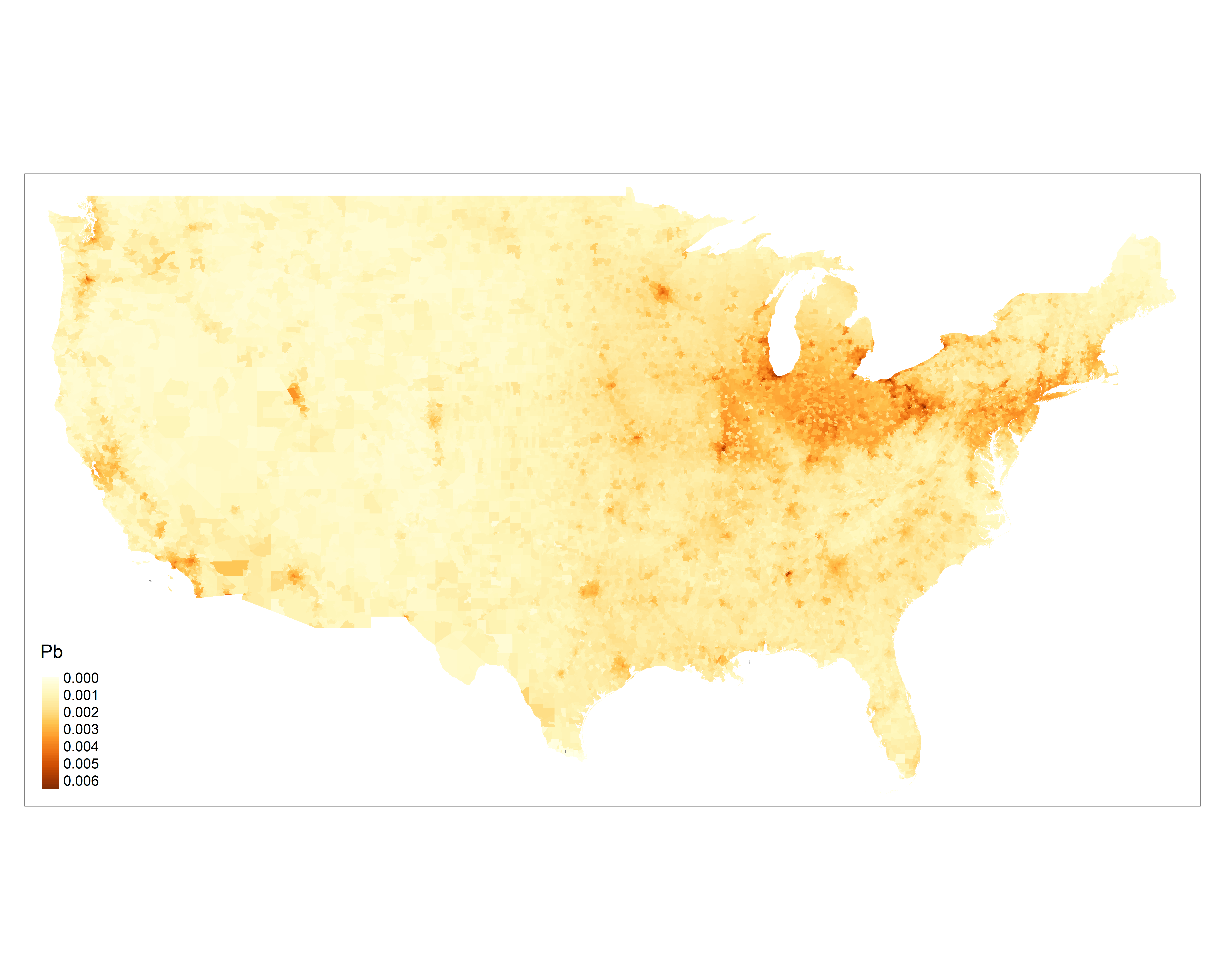

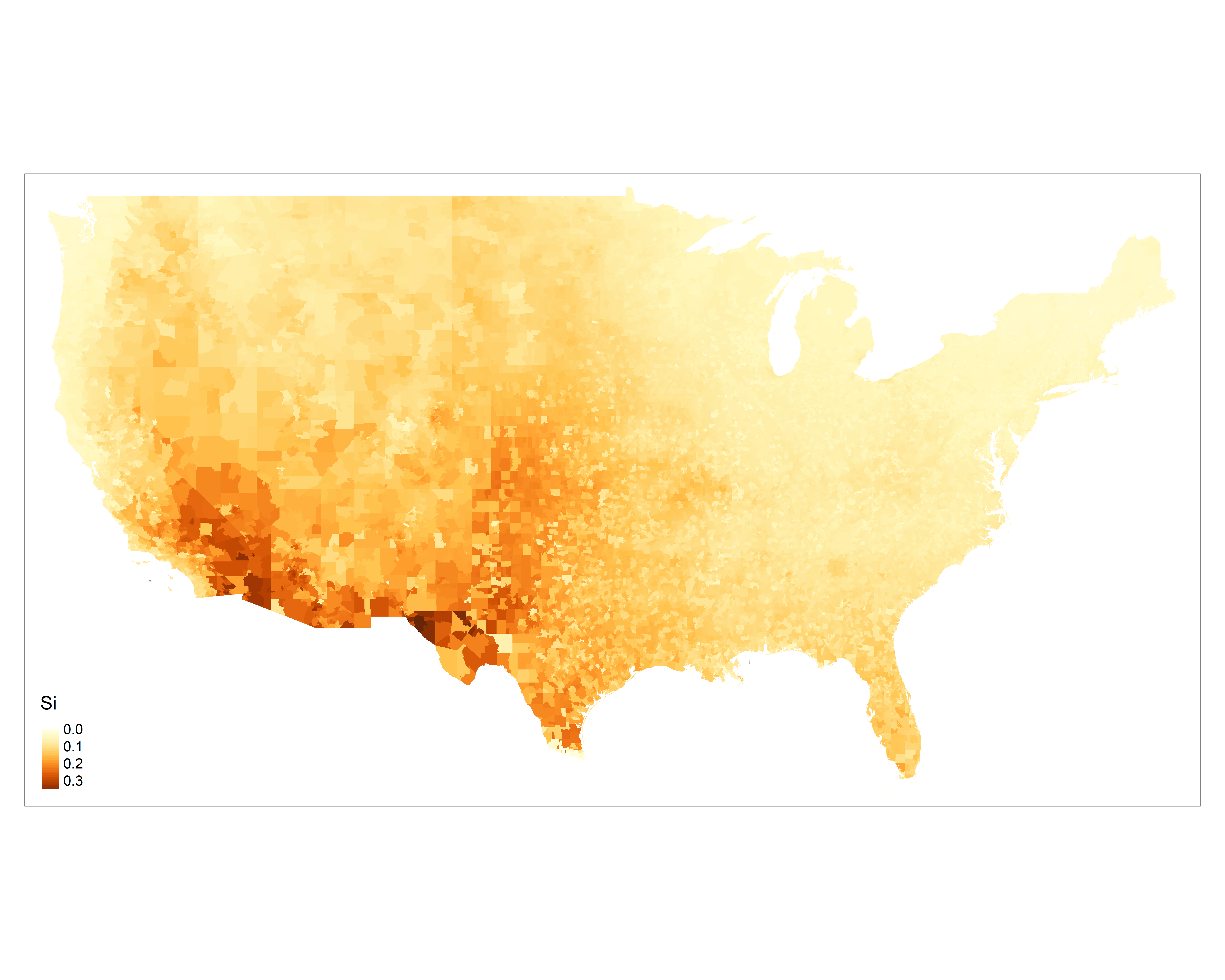

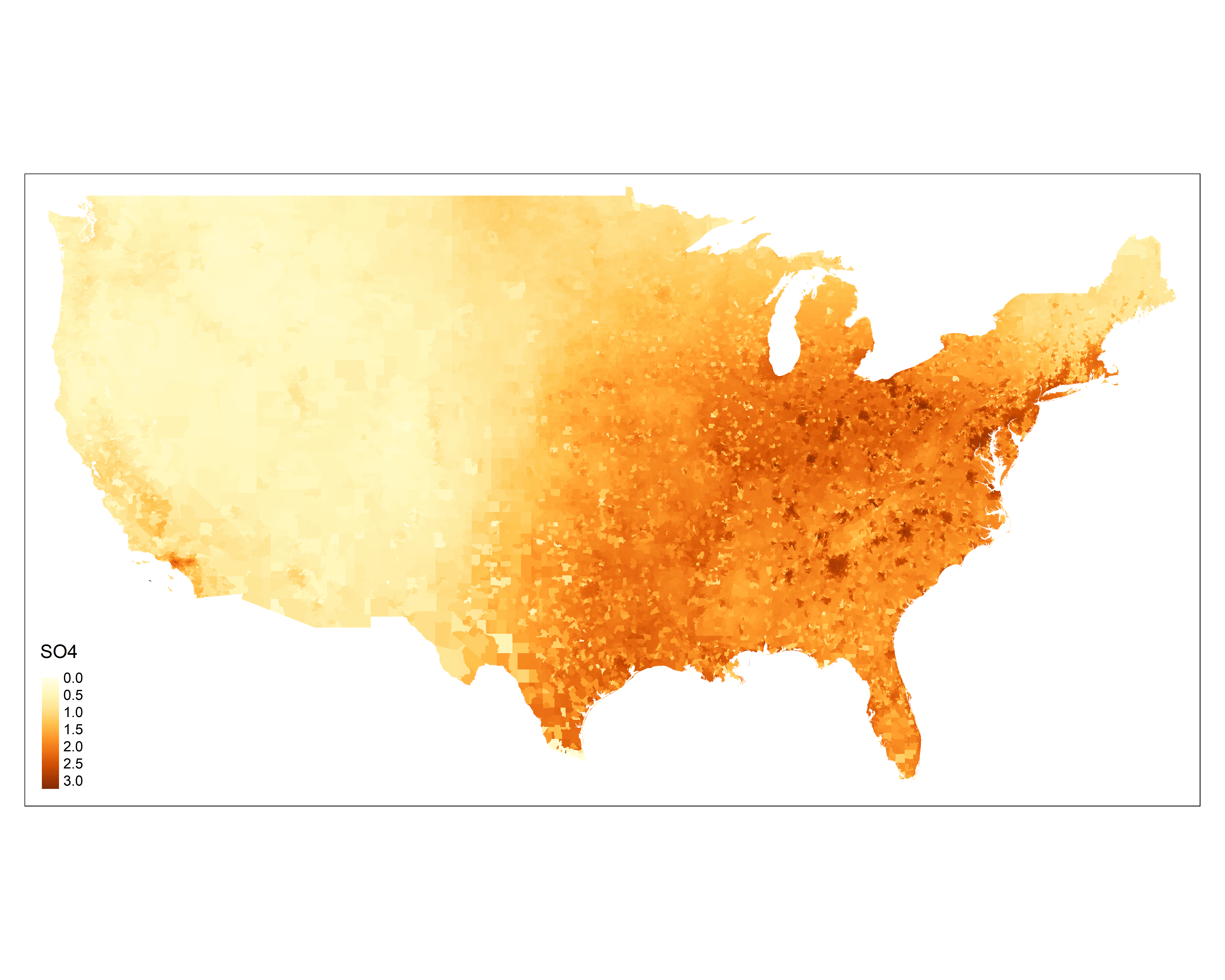

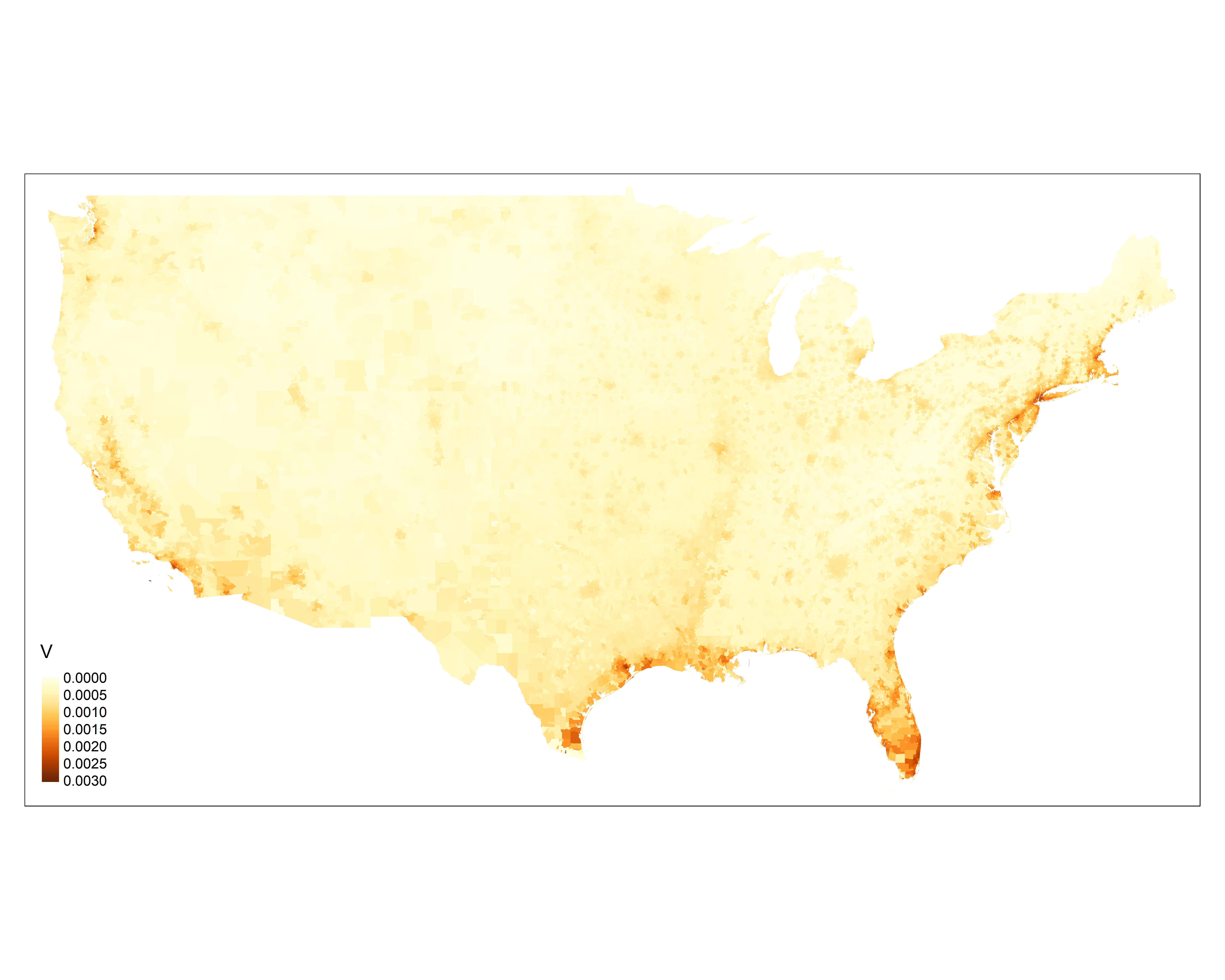

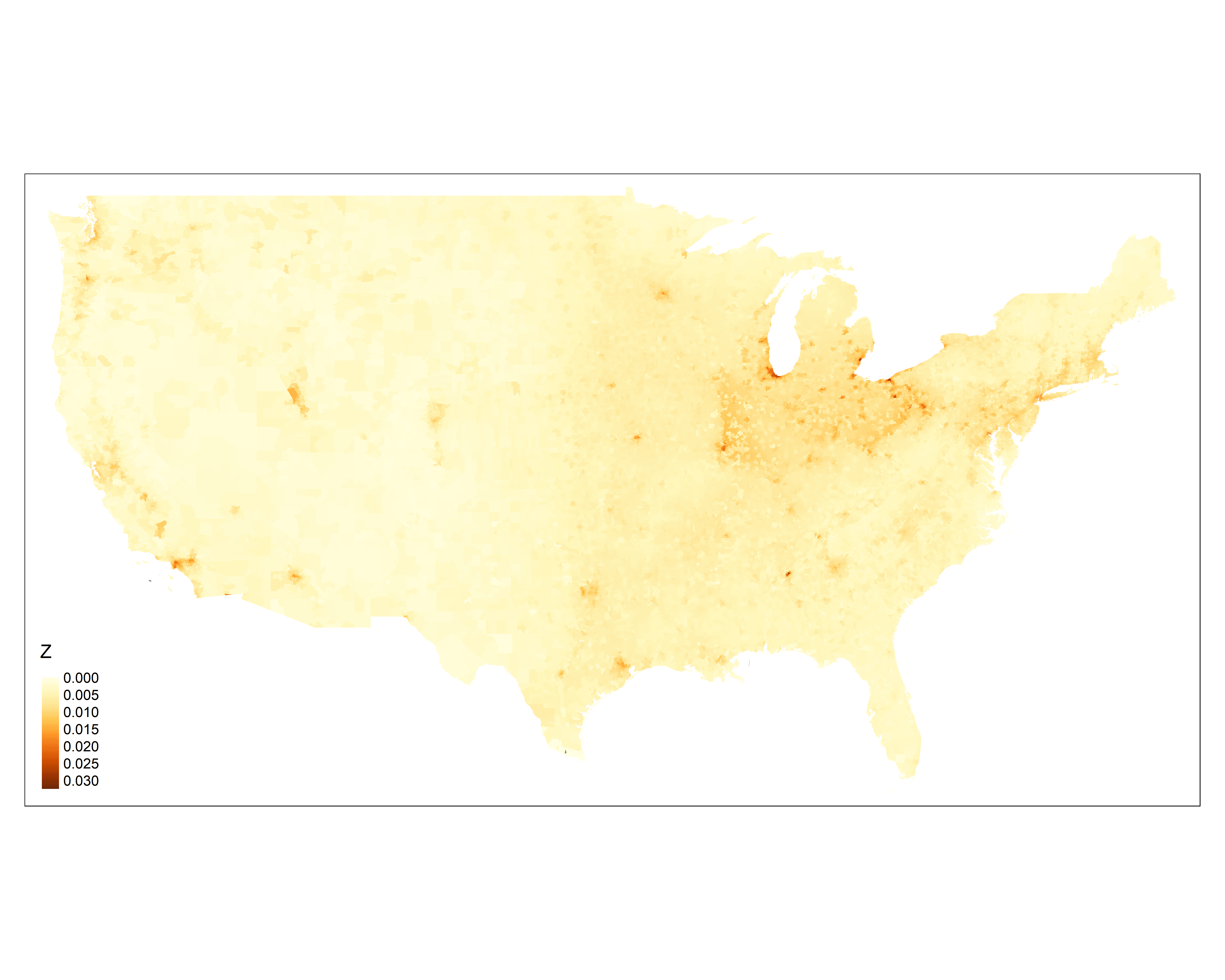

Figure S2: The correlation matrix among PM_2.5_ mass and 15 PM_2.5_ components. Pearson correlation coefficients were reported here.

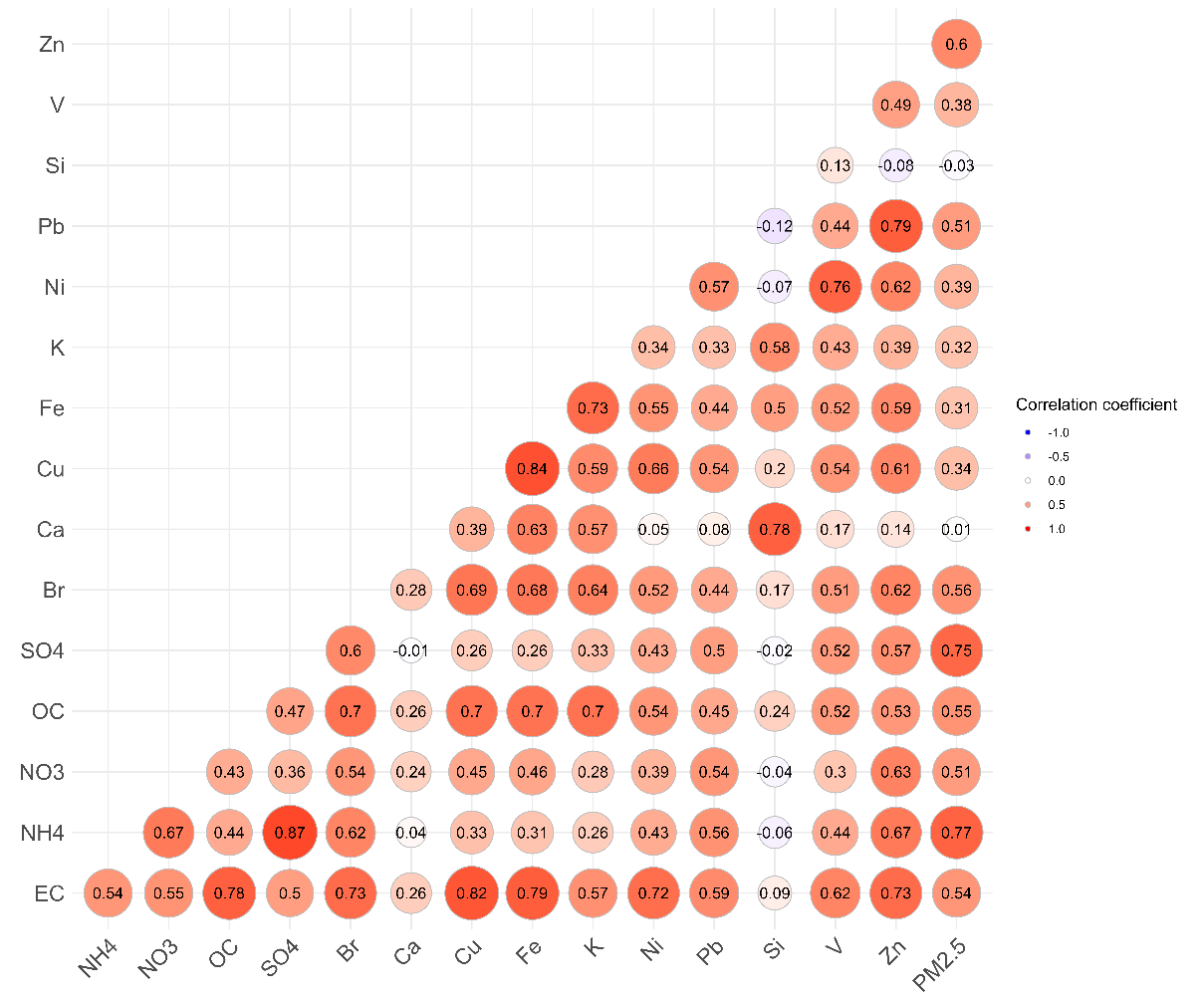

Figure S3: The annual mean concentrations of PM_2.5_ mass and PM_2.5_ components. The unit for concentrations are ug/m^3^ for major mass contributors (EC, OC, NH_4_^+^, NO_3_^-^, SO4^2-^) and PM_2.5_ mass; and pg/m^3^ for trace elements (Br, Ca, Cu, Fe, K, Ni, Pb, Si, V, Zn)

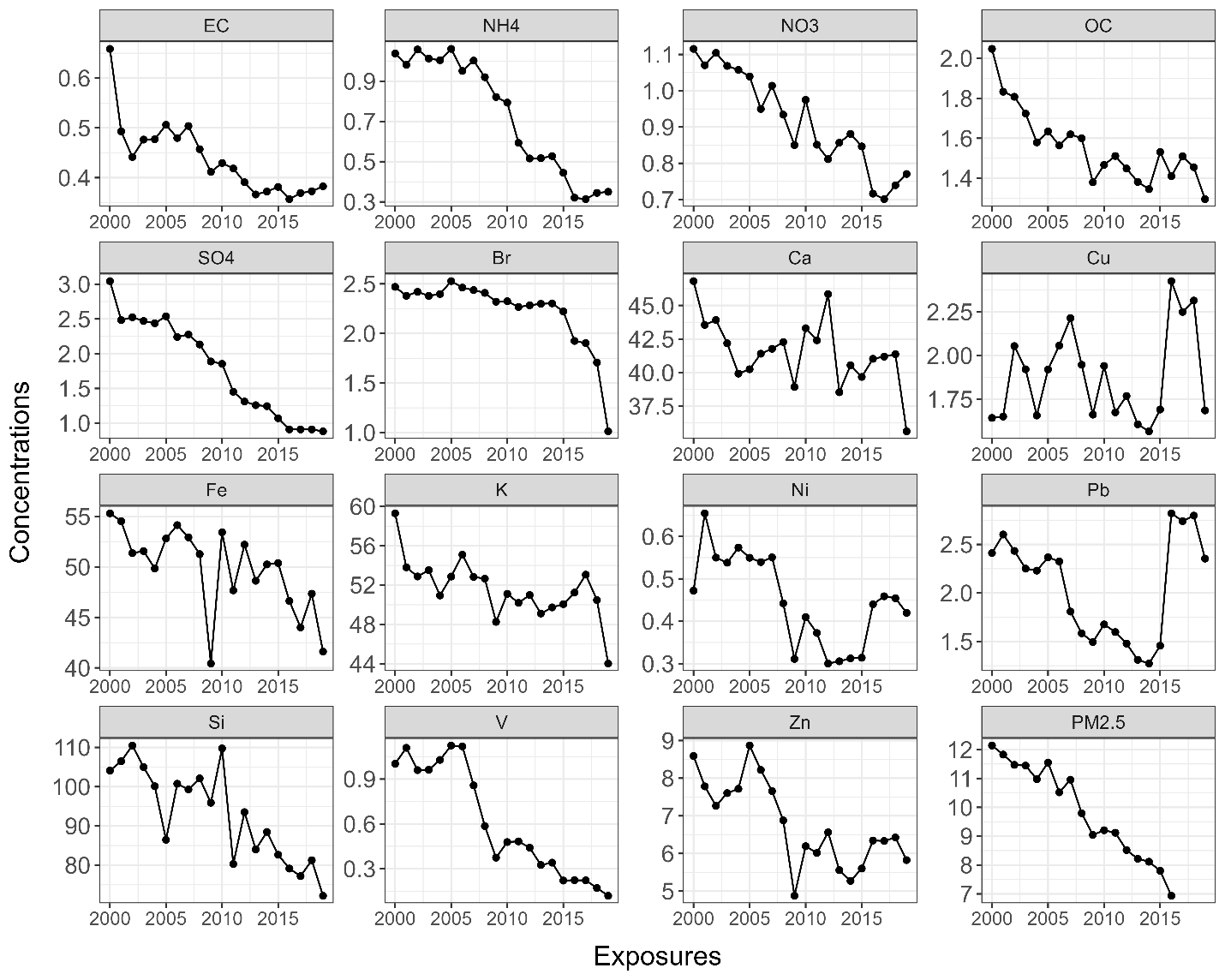

Figure S4: Hazard ratios (HRs) along with the 95% confidence intervals of dementia or Alzheimer’s disease associated with per IQR increase in annual mean concentrations of PM_2.5_ mass and PM_2.5_ components. HRs were estimated through single pollutant models.

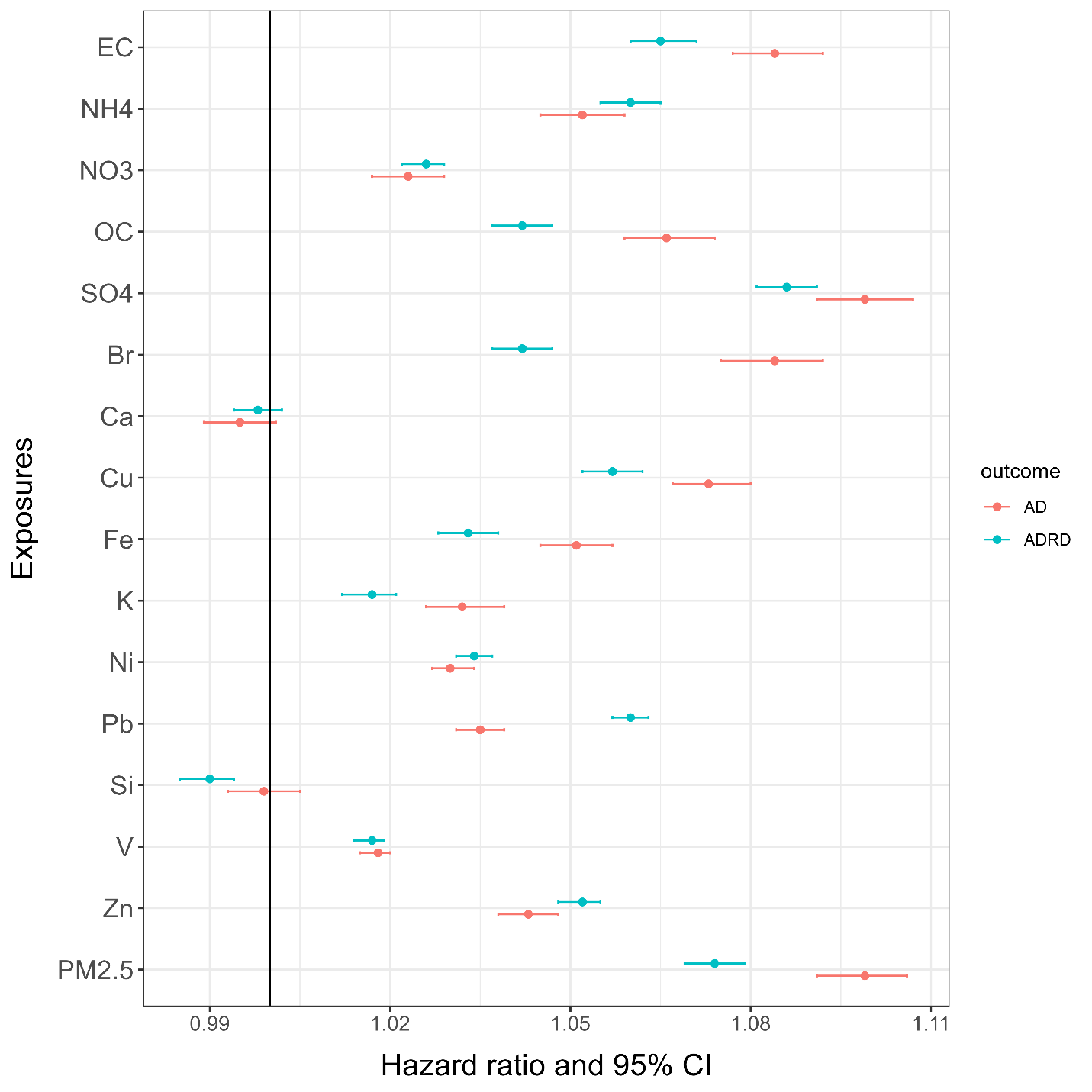

Figure S5: Hazard ratios (HRs) along with the 95% confidence intervals of dementia or Alzheimer’s disease associated with per IQR increase in annual mean concentrations of PM_2.5_ components. HRs were estimated through multi-pollutant models.

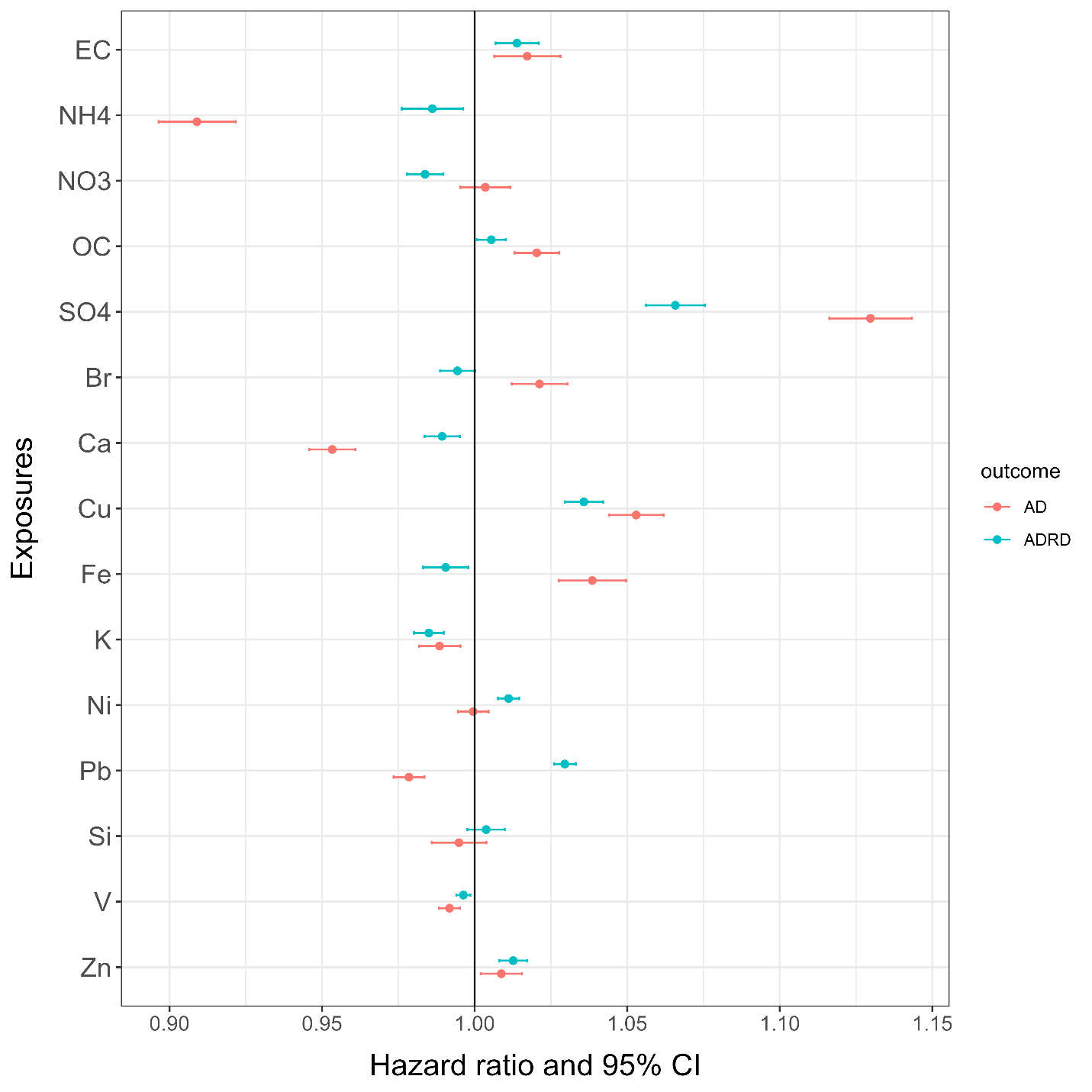

Figure S6: The weights assigned to each PM_2.5_ component from both WQS and qgcomp models among non-movers.

Non-movers:

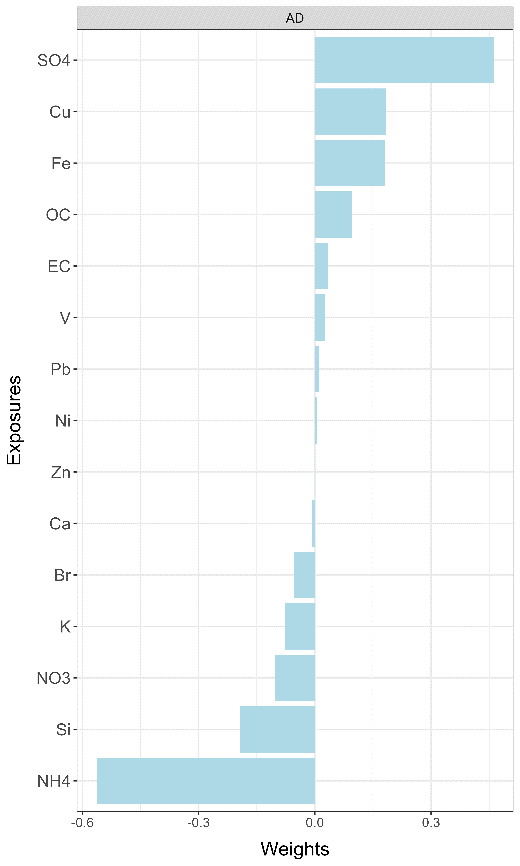

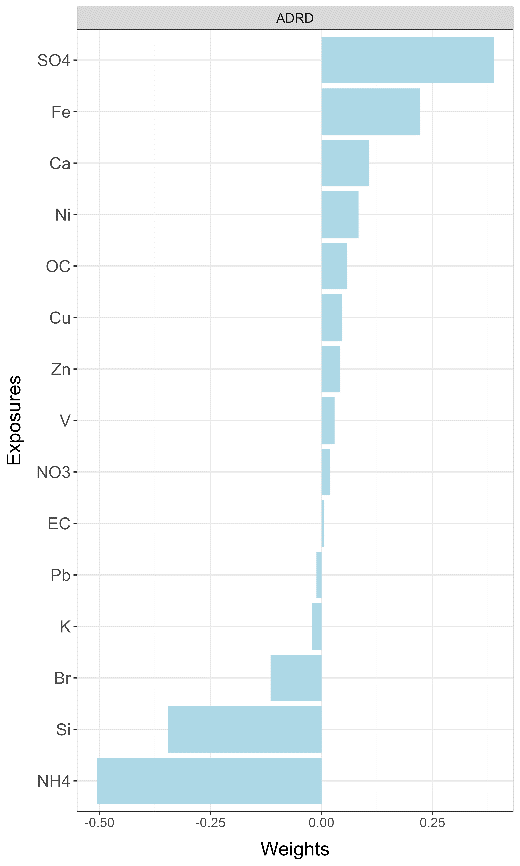

Non-movers:

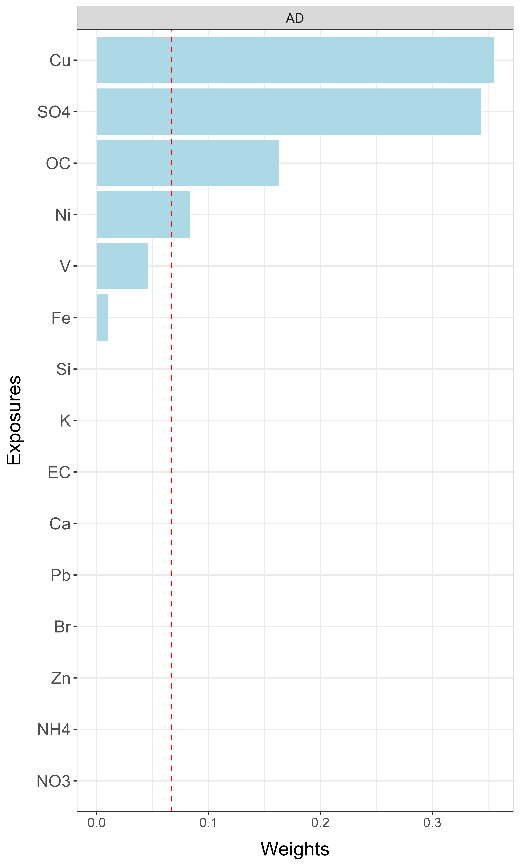

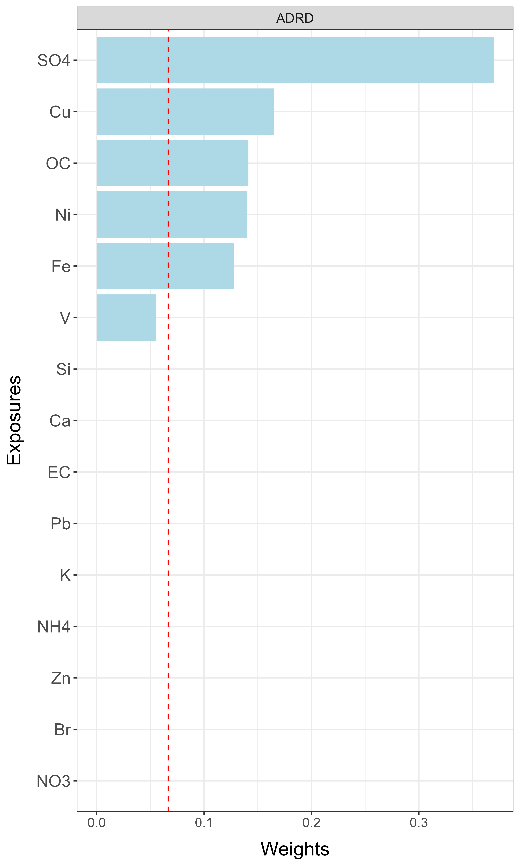

Figure S7: The weights assigned to each PM_2.5_ component from both WQS and qgcomp models with 3-year clean period and 4-year (same year and up to 3 years before) moving average exposures.

Lag:

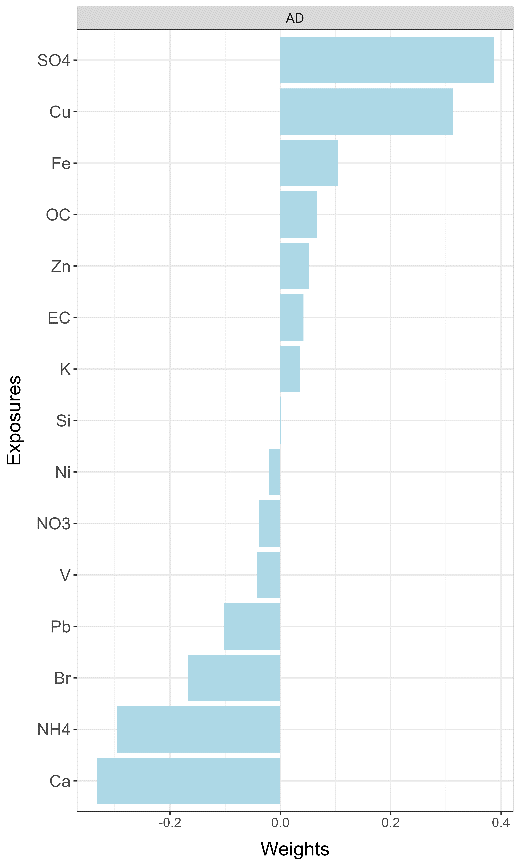

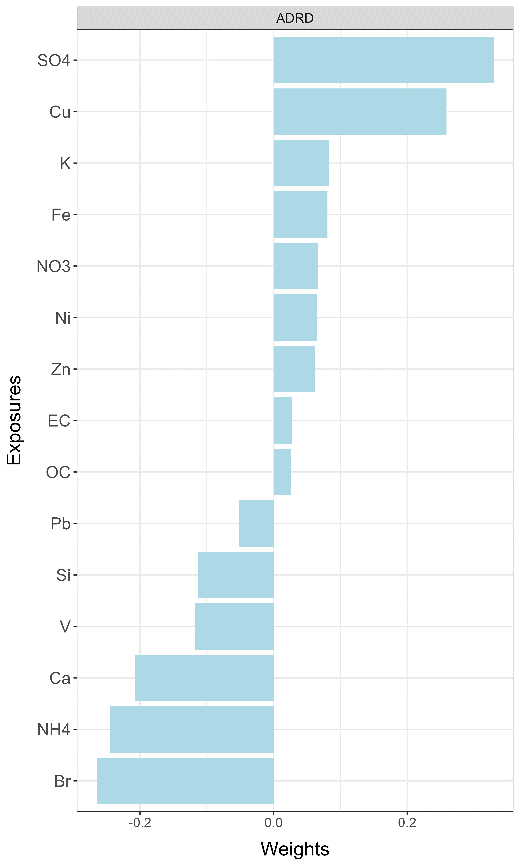

Lag:

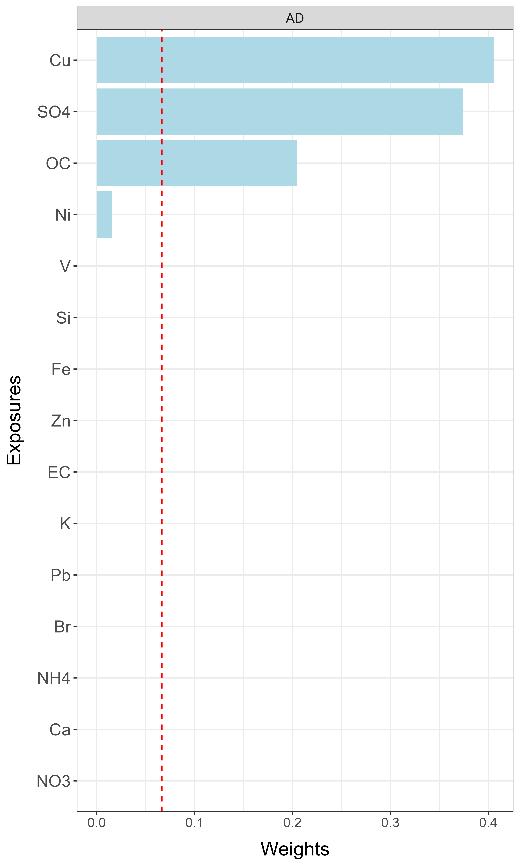

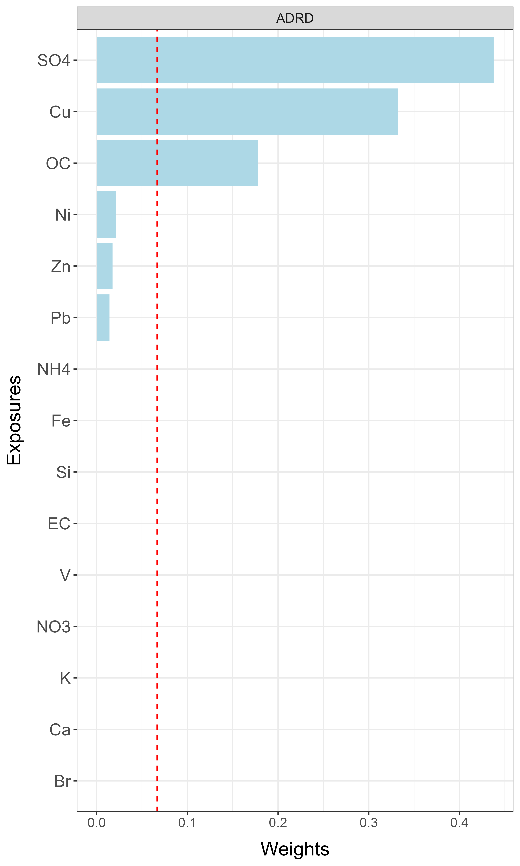

Table S1: ICD code associated with dementia / AD claim.

|  | AD | Dementia |
| --- | --- | --- |
| ICD-9 | DX 331.0 (any DX on the claim) | DX 331.0, 331.11, 331.19, 331.2, 331.7, 290.0, 290.10, 290.11, 290.12, 290.13, 290.20, 290.21, 290.3, 290.40, 290.41, 290.42, 290.43, 294.0, 294.10, 294.11, 294.20, 294.21, 294.8, 797 (any DX on the claim) |
| ICD-10 | DX G30.0, G30.1, G30.8, G30.9 (any DX on the claim) | DX F01.50, F01.51, F02.80, F02.81, F03.90, F03.91, F04, G13.2, G13.8, F05, F06.1, F06.8, G30.0, G30.1, G30.8, G30.9, G31.1, G31.2, G31.01, G31.09, G91.4, G94, R41.81, R54 (any DX on the claim) |

Table S2: The weights assigned to each PM_2.5_ component from WQS and qgcomp models.

|  | Alzheimer’s diseases | | Dementia | |
| --- | --- | --- | --- | --- |
|  | qgcomp | WQS | qgcomp | WQS |
| EC | 0.050 | 0 | 0.024 | 0 |
| NH4 | -0.310 | 0 | -0.216 | 0 |
| NO3 | -0.121 | 0 | 0.015 | 0 |
| OC | 0.091 | 0.221 | 0.047 | 0.24 |
| SO4 | 0.406 | 0.367 | 0.328 | 0.44 |
| Br | -0.193 | 0 | -0.281 | 0 |
| Ca | -0.226 | 0 | -0.078 | 0 |
| Cu | 0.312 | 0.405 | 0.265 | 0.264 |
| Fe | 0.080 | 0 | 0.074 | 0 |
| K | 0.016 | 0 | 0.084 | 0 |
| Ni | -0.032 | 0.006 | 0.067 | 0.055 |
| Pb | -0.020 | 0 | -0.091 | 0 |
| Si | -0.074 | 0 | -0.222 | 0 |
| V | -0.024 | 0 | -0.112 | 0 |
| Zn | 0.045 | 0 | 0.095 | 0 |

Table S3: Hazard ratios (HRs) along with the 95% confidence intervals of dementia or Alzheimer’s disease associated with per IQR increase in annual mean concentrations of PM_2.5_ components, from both single pollutant and multi-pollutant cox models.

|  | Single pollutant | | Multi-pollutant | |
| --- | --- | --- | --- | --- |
|  | ADRD | AD | ADRD | AD |
| EC | 1.065 (1.060, 1.071) | 1.084 (1.077, 1.092) | 1.014 (1.007, 1.021) | 1.017 (1.006, 1.028) |
| NH4 | 1.060 (1.055, 1.065) | 1.052 (1.045, 1.059) | 0.986 (0.976, 0.996) | 0.909 (0.896, 0.922) |
| NO3 | 1.026 (1.022, 1.029) | 1.023 (1.017, 1.029) | 0.984 (0.978, 0.990) | 1.003 (0.995, 1.012) |
| OC | 1.042 (1.037, 1.047) | 1.066 (1.059, 1.074) | 1.005 (1.001, 1.010) | 1.020 (1.013, 1.028) |
| SO4 | 1.086 (1.081, 1.091) | 1.099 (1.091, 1.107) | 1.066 (1.056, 1.075) | 1.130 (1.116, 1.143) |
| Br | 1.042 (1.037, 1.047) | 1.084 (1.075, 1.092) | 0.994 (0.989, 1.000) | 1.021 (1.012, 1.030) |
| Ca | 0.998 (0.994, 1.002) | 0.995 (0.989, 1.001) | 0.989 (0.984, 0.995) | 0.953 (0.946, 0.961) |
| Cu | 1.057 (1.052, 1.062) | 1.073 (1.067, 1.080) | 1.036 (1.030, 1.042) | 1.053 (1.044, 1.062) |
| Fe | 1.033 (1.028, 1.038) | 1.051 (1.045, 1.057) | 0.990 (0.983, 0.998) | 1.039 (1.028, 1.050) |
| K | 1.017 (1.012, 1.021) | 1.032 (1.026, 1.039) | 0.985 (0.980, 0.990) | 0.989 (0.982, 0.995) |
| Ni | 1.034 (1.031, 1.037) | 1.030 (1.027, 1.034) | 1.011 (1.008, 1.015) | 1.000 (0.995, 1.005) |
| Pb | 1.060 (1.057, 1.063) | 1.035 (1.031, 1.039) | 1.030 (1.026, 1.033) | 0.978 (0.973, 0.984) |
| Si | 0.990 (0.985, 0.994) | 0.999 (0.993, 1.005) | 1.004 (0.998, 1.010) | 0.995 (0.986, 1.004) |
| V | 1.017 (1.014, 1.019) | 1.018 (1.015, 1.020) | 0.996 (0.994, 0.999) | 0.992 (0.988, 0.995) |
| Zn | 1.052 (1.048, 1.055) | 1.043 (1.038, 1.048) | 1.013 (1.008, 1.017) | 1.009 (1.002, 1.016) |
| PM_2.5_ | 1.074 (1.069, 1.079) | 1.099 (1.091, 1.106) |  |  |

Table S4: Cumulative associations between PM2.5 components and Alzheimer’s diseases / dementia in rate ratios estimated from both WQS and qgcomp models under different settings.

| Models | Mover status | Clean & lag period | AD | ADRD |
| --- | --- | --- | --- | --- |
| qgcomp |  |  |  |  |
|  | All | 5yr | 1.041 (1.039, 1.042) | 1.027 (1.025, 1.028) |
|  | Non-movers | 5yr | 1.036 (1.034, 1.038) | 1.022 (1.020, 1.023) |
|  | All | 3yr | 1.041 (1.039, 1.042) | 1.027 (1.025, 1.028) |
| WQS |  |  |  |  |
|  | All | 5yr | 1.048 (1.047, 1.049) | 1.029 (1.028, 1.030) |
|  | Non-movers | 5yr | 1.048 (1.046, 1.049) | 1.027 (1.026, 1.028) |
|  | All | 3yr | 1.049 (1.047, 1.050) | 1.031 (1.031, 1.032) |
